# MR-link-2: pleiotropy robust *cis* Mendelian randomization validated in four independent gold-standard datasets of causality

**DOI:** 10.1101/2024.01.22.24301400

**Authors:** Adriaan van der Graaf, Robert Warmerdam, Chiara Auwerx, eQTLGen Consortium, Urmo Võsa, Maria Carolina Borges, Lude Franke, Zoltán Kutalik

## Abstract

Mendelian randomization (MR) can identify causal relationships from observational data but has increased Type 1 error rates (T1E) when genetic instruments are limited to a single associated region, a typical scenario for molecular exposures. To address this, we developed MR-link-2, which uses summary statistics and linkage disequilibrium (LD) information to simultaneously estimate a causal effect and pleiotropy in a single associated region. We extensively compare MR-link-2 to other *cis* MR methods: i) In realistic simulations, MR-link-2 has calibrated T1E and high power. ii) We replicate causal relationships derived from three metabolic pathway references using four independent metabolite quantitative trait locus studies as input to MR. Compared to other methods, MR-link-2 has a superior area under the receiver operator characteristic curve (AUC) (up to 0.80). iii) Applied to canonical causal relationships between complex traits, MR-link-2 has a lower per-locus T1E rate than competing methods (0.09 vs 0.15, at a nominal 5% level) and has several fold less heterogeneous causal effect estimates. iv) Testing the correct causal direction between blood cell type compositions and gene expression of their marker genes reveals that MR-link has superior AUC 0.90 (best competing: 0.67). Finally, when testing for causality between metabolites that are not connected by canonical reactions, MR-link-2 exclusively identifies a link between glycine and pyrroline-5-carboxylate, corroborating results for hypomyelinating leukodystrophy-10, otherwise only found in model systems. Overall, MR-link-2 is the first method to identify pleiotropy-robust causality from summary statistics in single associated regions, making it ideally suited for applications on molecular phenotypes.

## Main Text

The identification of causal relationships in humans is historically done using randomized control trials (RCTs). However, these trials require the separation of individuals into treatment and control groups, which is a burden on the subjects and can be expensive. Additionally, some trials cannot be carried out due to ethical considerations or are simply impossible to perform as there is no suitable way to perturb the trait under investigation. Observational causal inference attempts to identify causal relationships from observational data by identifying randomization events that occurred naturally in a group of individuals. If performed correctly, observational causal inference can be useful in improving our understanding of the causal relationships that underlie human biology, aiding in the development or repurposing of drugs and treatments. One observational causal inference technique that is popular in the genetics community is Mendelian randomization (MR) (*1, 2*). For instance, MR has shown to be effective in identifying the causal relationship between low density lipoprotein cholesterol (LDL-C) levels and coronary heart disease and between alcohol consumption and cardiovascular disease (*3, 4*), while careful application of MR has shown that high density lipoprotein cholesterol (HDL-C) levels are not causally linked to myocardial infarction, evidence that was also corroborated by RCTs (*5, 6*). A valid MR analysis is done based on three statistical assumptions: i) The relevance assumption, ii) the independence assumption and iii) the exclusion restriction (also known as pleiotropy) (**Fig. 1a**), described in more detail in (**Supplementary Text**). Considering the assumptions underlying MR, it is particularly difficult to ensure that the genetic variants that are used as instrumental variables (IVs) are free from horizontal pleiotropy. As it is usually impossible to ensure that a genetic variant only acts through the chosen exposure (*7*). MR methods are constantly being developed to ensure MR estimates are robust (*8*–*13*). Generally, these methods use tens to hundreds of independent locations on the genome in a meta-analysis to mitigate violations of the relevance assumption and exclusion restriction with the hope that independent instruments would lead to independent biases which cancel each other out. However, when considering molecular traits, such as RNA and protein expression or metabolite concentrations, there is generally only a single or a handful of associated regions, reducing the robustness of these ‘meta-analyzing’ MR methods. Improving MR methods for application on molecular exposures is of high priority as these exposures are important causes for disease and are potential drug-targets. Currently, identification of molecular traits as causes to disease often relies on “closest gene analysis”, tools like MAGMA and PASCAL or colocalization methods (*14*–*18*). These approaches have shown that they can identify the correct molecule, but they do not strictly test for causality. For instance, a closest gene analysis will not identify the context in which a gene has its effect while colocalization analysis will only indicate if two traits share the same causal genetic variant(s). It is not answering the question if one is causal to the other or if there is a shared causal confounder. Furthermore, if a molecule under investigation has more than one associated genetic locus, it is difficult to combine the information into a single estimate of relevance. In contrast, MR has the benefit that it identifies a causal relationship, possibly from multiple loci. Unfortunately, except for some obvious examples, we lack good sources of truth for true causal links and false causal links between (molecular) traits in humans (*9, 19*). This limits our ability to compare different MR methods, as methodologists usually use simulations and single examples to highlight the strength of their causal inference method.

**Fig. 1.**
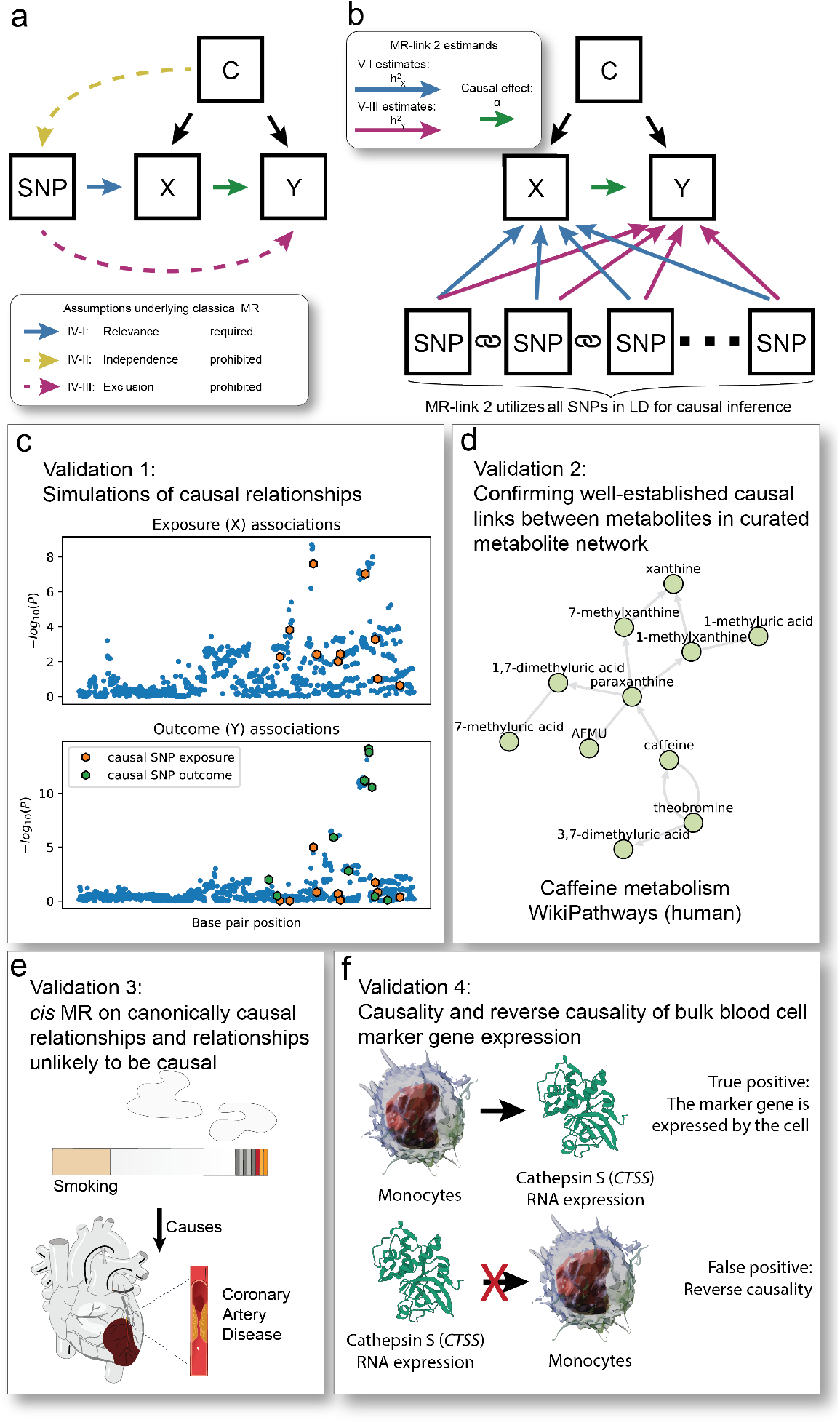
Overview of this study: the assumptions underlying Mendelian randomization (MR), a graphical representation of MR-link-2 method and the four ways we benchmark and compare MR-link-2 to other *cis* MR methods. (**a**) Directed acyclic graph to illustrate the assumptions underlying MR. Single nucleotide polymorphisms (SNPs) are used as instruments to estimate the causal effect between an exposure (X) and an outcome (Y) confounded by C. The blue, yellow and purple arrows highlight the assumptions underlying MR. Black arrows are allowed but are not necessary for correct inference. (**b**) Graphical representation of the MR-link-2 method. In contrast to other MR methods, MR-link-2 models all the SNPs in a genetic region to simultaneously estimate the (local) *cis* heritability of the exposure (IV-I, 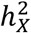, blue arrows), the total pleiotropic effects on the outcome due to violations of the exclusion restriction assumption (IV-III, 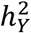, purple arrows) and the causal effect *α* (green arrow) that is robust to violations of IV-III. MR-link-2 requires that linkage disequilibrium is measured in between the genetic variants (chain symbol). (**c**-**f**) Validations done to compare MR-link-2 to other methods. (**c**) First validation is done using simulations. Shown here is a simulated genetic region where an exposure is causal to an outcome. The outcome also contains genetic effects independent of the exposure, which would violate the exclusion restriction (IV-III). (**d**) We perform a second comparison of *cis* MR methods using gold standard metabolite reactions present in curated metabolic networks. For illustration, we show here the human caffeine metabolism from WikiPathways. (**e**) Validation through canonical causal relationships between complex traits. Shown here, for illustration, is the well-known causal relationship between smoking and coronary artery disease. (**f**) Final validation tests the ability to decide between forward *vs* reverse causal effects. We utilize the genetics of blood cell proportions to predict their causal effect onto well-known blood cell marker genes. Null causal effects are defined as the reverse direction which should not be causal.

This study has two goals: i) to introduce a novel summary statistics *cis* MR method that is robust to horizontal pleiotropy and ii) to develop gold-standard datasets for the validation of (*cis*) MR methods. First, we introduce a summary statistics MR method that is robust to pleiotropy even when only a single region is available for analysis, making it suitable for the analysis of molecular traits as risk factors. We coin the method “MR-link-2” (**Fig. 1b**). Conceptually, MR-link-2 uses the region surrounding the genetic variant around the IV to estimate the effects of pleiotropy (*20*). In contrast to the original MR-link (v1), MR-link-2 does not require individual level data but can be used with summary statistics of traits, allowing for more widespread applications (*20*). To our knowledge, MR-link-2 is the only summary statistics MR method that explicitly models pleiotropy between two traits when only a single associated genetic region is available. The second main novelty lies in the development of benchmarking datasets. We benchmark MR-link-2 against other *cis* MR methods as well as two colocalization methods using extensive simulations (**Fig. 1c**) (**Table 1**) and three real-world datasets of true and false causal links. In the first real data validation, we create a metabolite network using three sources of curated databases of human metabolite pathways and we assess discriminative performance of each method using metabolite quantitative trait loci (mQTLs) that are derived from four different studies (**Fig. 1d**). Second, we assess the performance of MR methods on known causal relationships between complex traits, as well as relationships that are unlikely to be causal (**Fig. 1e**). Third, using new data from the full *trans* mapping of gene expression by the eQTLGen Consortium, we test for the causal relationship between blood cell composition and whole-blood expression levels of cell-type specific marker genes (**Fig. 1f**) (*21*).

**Table 1.** Feature and requirement comparison of the traits used in this study. We compare 6 different methods for the identification of *cis* genetic regulation. We specify if meta-analysis of multiple regions is possible for each method, if the genetic method uses all genetic variants that are available in a genetic region, if the method requires an LD reference and if the method has been designed to handle horizontal pleiotropy. ^a^ MR-IVW needs LD information only to perform the pruning step.

| Method | Meta-analysis possible | Uses all SNPs in a region | Requires an LD reference | Robust to horizontal pleiotropy | Tests for |
| --- | --- | --- | --- | --- | --- |
| MR-link-2 | Yes | Yes | Yes | Yes | Causality between two traits and presence of horizontal pleiotropy in the associated region |
| MR-IVW | Yes | No | No<sup>a</sup> | No | Causality between two traits |
| MR-IVW-LD | Yes | No | Yes | No | Causality between two traits |
| MR-PCA | Yes | Yes | Yes | No | Causality between two traits |
| coloc | No | Yes | No | Yes | If a single causal SNP in a region is shared between two traits |
| coloc-SuSiE | No | Yes | Yes | Yes | If any of the causal genetic variants in a region are shared between two traits |

In all validation datasets, MR-link-2 compares favorably to other methods, exhibiting lower type 1 error (T1E) and good discriminative performance between true and false causal links, which we attribute to the method’s robustness to the presence of horizontal pleiotropy. When considering results outside of the validation datasets, MR-link-2 uniquely identifies regulation between metabolites as well as disease-relevant metabolite interactions that relate to hypomyelating leukodystrophy 10 (HDL10).

### The MR-link-2 method

MR-link-2 is a likelihood function that estimates three parameters based on the exposure and the outcome summary statistics in a region, combined with a reference linkage disequilibrium (LD) matrix (**Fig. 1b**) (**Methods**). MR-link-2 tests for two parameters using a likelihood ratio test: the causal effect estimate 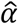, which is of central interest, and the remaining horizontal pleiotropic variance, 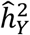,which would otherwise violate the exclusion restriction (**Methods**) (**Supplementary Text**). The modeling of the pleiotropic variance allows MR-link-2 to be robust to violations of the pleiotropy assumption using only a single genetically associated region. We designed MR-link-2 to work in scenarios that we find biologically plausible, however, as with any statistical method, MR-link-2 relies on some modeling assumptions. The likelihood function of MR-link-2 can be sensitive to three parameters: i) when the LD matrix is measured with imprecision (either due to small sample size of the reference panel or population mismatch), ii) when there is a large amount of LD between underlying causal SNPs and iii) when there is a small number of causal SNPs underlying the exposure and the outcome trait.

### Simulations

We performed simulations of causality in a single genetic region with two goals: first, to understand the statistical behavior of MR-link-2 when the parameters in the simulations are varied, including when assumptions underlying MR (**Fig. 1a**) and MR-link-2 are violated. Secondly, to compare the performance of MR-link-2 with other *cis* MR methods (**Methods**). Across 2,700 simulation parameter settings, we simulated 1,000 exposure and outcome pairs that are genetically regulated by a single region based on LD derived from the UK10K cohort (*22*).

We varied 6 parameters: the simulated causal effect (*α*), the *cis* heritability of the exposure 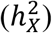 and the extent of pleiotropy of the outcome 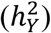, the number of causal SNPs underlying both the exposure and the outcome (*m*_*casual*_), their minimum correlation between the causal markers for the exposure and those with direct causal effect on the outcome (min (*r*_*casual*_)) and imprecision in the LD reference (parameterized by the reference panel size, *n*_*ref*_) (**Methods**).

In simulations with 100 causal exposure and outcome SNPs and an LD matrix measured with full precision, MR-link-2 has well calibrated T1E (min: 0.01, median 0.05, max 0.07), across the range of simulated exposure heritabilities including when there is strong violation of the pleiotropy assumption (**Fig. 2a**) (**Data S1**). When simulating a large causal effect 0.2 (**Fig. 2b**) (**Data S1**), we found that increasing the exposure genetic variance increased detection power (up to 1.00), whereas increasing pleiotropy reduced detection power **(Fig. 2b)**. T1E rates generally increased when simulating violations in the MR-link-2 assumptions. MR-link-2 has increased T1E rates when we introduce imprecision in the LD reference (up to 0.42 when the LD reference is measured only in 500 individuals) (**Data S1**), when causal genetic variants of the two traits are in very strong LD (up to 0.243 when SNPs are in LD *r*^2^ > 0.1 to each other) (**Data S1**). However, MR-link-2 is not dependent on the number of causal SNPs that underlie a trait (max T1E rate: 0.05 when simulating 1 causal SNP for both traits) (**Data S1**). When violating all these assumptions together, the T1E rate increased to 0.84 when simulating a single causal SNP combined with an extremely large 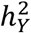 of 0.03 (**Data S1**), even though this situation is unlikely to occur in human biology, as it is highly unusual to find single variants with such a large effect on complex outcomes. A unique feature of MR-link-2 is that it can identify residual genetic variance in the outcome, which would otherwise be modeled as violations of the exclusion restriction. When simulating minute 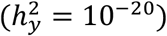 pleiotropy and following the MR-link2 underlying assumptions, MR-link-2 does not detect pleiotropy (detection rate minimum: 0.00, maximum: 0.01) with deflated test statistics compared to the expected 0.05 (**Fig. 2c**) (**Data S1**). However, when simulated pleiotropy is increased above 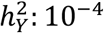in the simulation, MR-link-2 correctly estimates the extent of pleiotropy (**Fig. 2c**). To ensure that MR-link-2 is adequately powered and has good discriminative ability, we compared MR-link-2 to three other (*cis*) MR methods (MR-IVW, MR-IVW LD and MR-PCA) as well as to two colocalization methods (coloc and coloc SuSiE) using the area under the receiver operator characteristic curve (AUC) metric (*14, 15, 23*–*25*) (**Table 1**) (**Fig. 2d-h**) (**Data S2**). In many cases, we find that the AUC of MR-link-2 is higher, especially when simulating pleiotropy 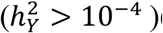 (**Fig. 2f**). To understand the influence of each parameter setting on the discriminative ability of each method, we performed ordinary least squares regression with all model parameters as predictors and the AUC of a method as the dependent variable (**Fig. 2i**) (**Data S3**). Here we see that the AUC generally decreased for each method as pleiotropy is simulated, with the smallest decline observed for MR-link-2, providing further evidence for robustness to pleiotropy of our method (**Fig. 2i**) (**Data S3**). Furthermore, we see that the imprecision of the reference panel negatively influences only MR-link-2 and coloc SuSiE (**Fig. 2i**).

**Fig. 2.**
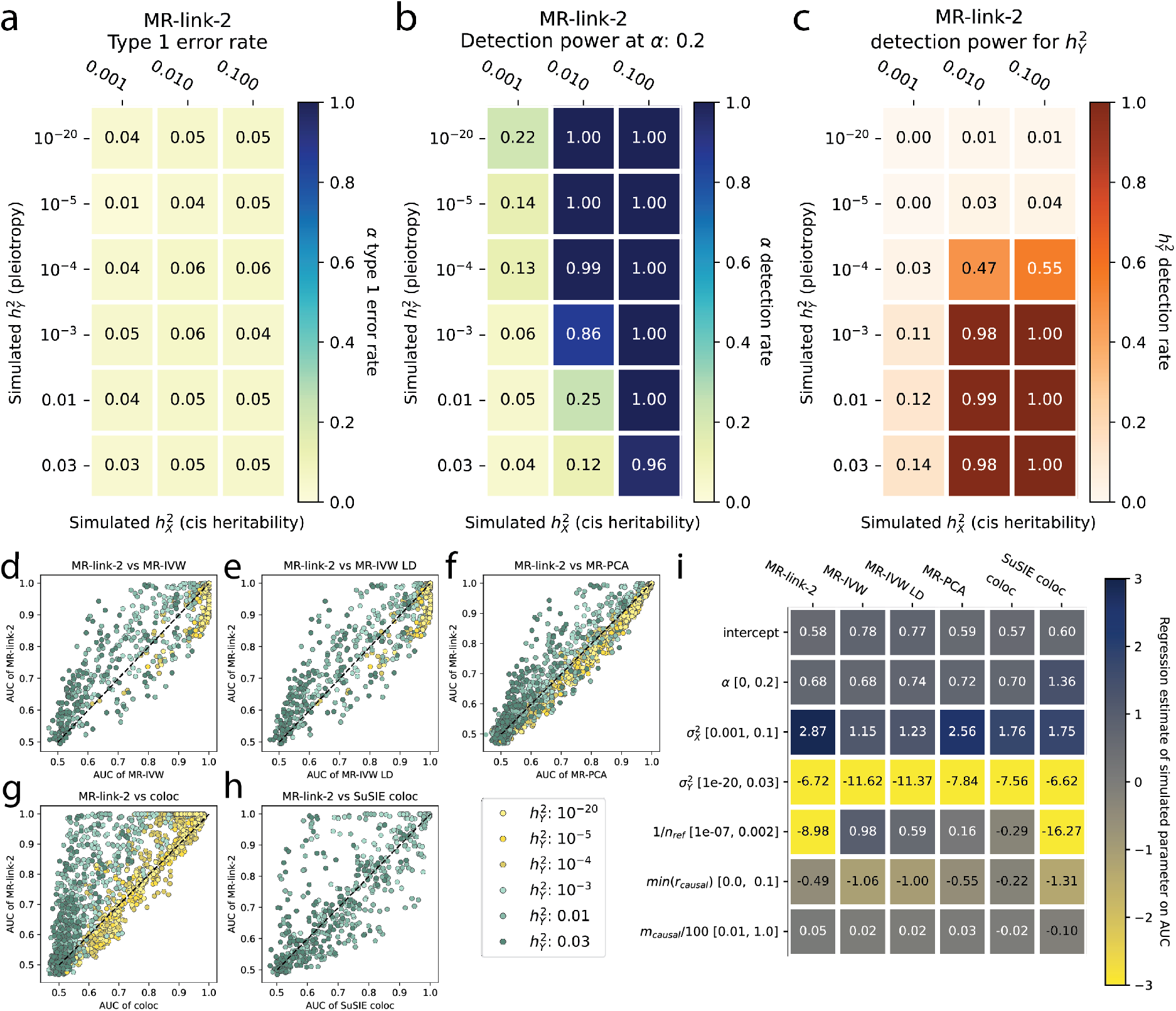
Simulations of MR-link-2 in different scenarios. (**a**) Type I error rate of MR-link-2 in simulations with no causal effect (*α* = 0) and various combinations of exposure genetic variance (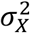, which is a measure of IV-I) and outcome genetic variance (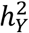,which violates the IV-III assumption of no pleiotropy). (**b**) Statistical power in the same simulation scenarios as panel (a) with a simulated causal effect (*α* = 0.2). (**c**) The power to detect non-zero pleiotropy by MR-link-2 (testing the pleiotropy parameter 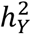). The simulation settings are the same as in panel (a), however, here we do not test for a causal effect, rather we test for violations of the IV-III assumptions of no pleiotropy. (**d-h**) The discriminative ability of MR-link-2 and other tested methods between simulations of no causal effect and those with a non-zero causal effect, characterized by the area under the receiver operator characteristic curve (AUC). The AUC values of MR-link-2 are compared to those of other competing methods. Here we also included additional simulation scenarios, where the infinitesimal exposure genetic model is violated (**Methods**). Parameter settings are only plotted for which both methods successfully estimate at least 750 / 1000 simulation instances in both null and non-null causal effect scenarios. Points are colored by the simulated pleiotropy parameter of 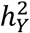. The x-axis corresponds to methods as follows: (**d**) MR-IVW; (**e**) MR-IVW LD; (**f**) MR-PCA; (**g**) coloc; (**h**) coloc SuSIE. (**Methods**) (**Data S2**) (**i**) A heatmap of (multivariable ordinary least squares) regression coefficients for each method when AUC is regressed on various model parameters. This allows identification of the impact of each simulation parameter on the AUC of each method. The simulated range of each parameter is shown in brackets. 1/*n*_*ref*_: represents the precision of the linkage disequilibrium reference used in this study, i.e. the inverse of the reference panel size. min (*r*_*casual*_) represents the minimum correlation between the causal SNPs and SNPs with direct effect on *Y*. - *m*_*casual*_*/*100 represents the number of causal SNPs selected in the region divided by 100 to ensure comparable regression coefficient scales (**Methods**).

### Using metabolite networks as a source of true causal links

Some causal relationships between complex traits are well known (*9*), however, when considering molecular phenotypes such as RNA expression, there are only a few examples of causal relationships that are known and which can be reliably tested through genetic methods (*19*). Leveraging canonical causal relationships from metabolite networks is a promising avenue since some of them have been known for more than 85 years and have been experimentally validated (*26*). We use these causal relationships as ground truth, to understand when MR methods fail and to subsequently compare MR methods.

Our ground truth metabolite network is derived from the human metabolic pathway definitions of KEGG, MetaCyc and WikiPathways (*27*–*29*) (**Fig. 3a**) (**Methods**). Orthogonally, we applied MR/colocalization methods to four mQTL studies comprising of 1,291 harmonized metabolite measurements of 1,035 unique metabolites (*30*–*33*) (**Methods**) (**Fig. 3a**). After harmonization with the pathway definitions, we kept 266 measurements across mQTL studies, representing 154 unique metabolites. One hundred ninety-three metabolite measurements have an mQTL at *P ≤* 5 ⋅ 10^−8^, representing 126 separate metabolites which can be used as exposures to compare their causal effects to the “ground truth” (**Fig. 3a-c**). Across these 154 unique metabolites, our pathway definitions define 287 individual chemical reactions that can be used as true causal links. (**Fig. 3a-c**) (**Methods**) (**Data S4**). Comparing pathway definitions between each other, the concordance of MetaCyc and WikiPathways was the highest, while KEGG was less concordant. Indeed, only 34 out of 284 reactions are present in all three pathway databases, which may be due to differing curation standards. Fifty-five reactions are shared by at least two pathway databases and the remaining 194 reactions are specific to single pathway databases (**Fig. 3c**).

**Fig. 3.**
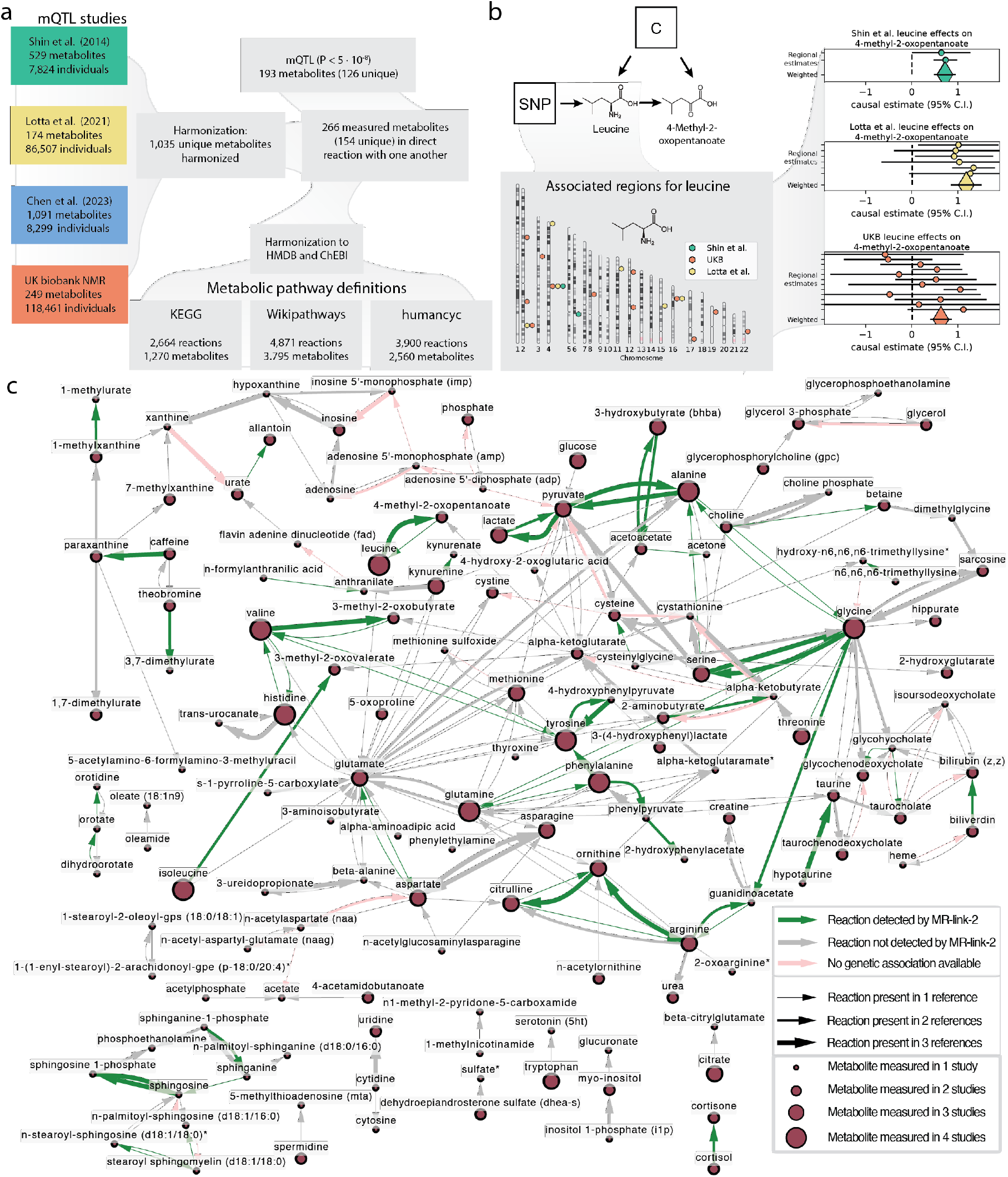
Metabolite quantitative trait loci (mQTL) studies used in this analysis, an example MR analysis and the true causal links and true positives identified in this study. **(a)**Chart depicting the metabolites and their QTLs used in this study. We utilized four mQTL studies whose studied metabolites were harmonized into 1,037 consensus metabolites. To create ground truth causal links between these metabolites, we used three pathway definitions. Overlapping the measured metabolites from the mQTL studies with the metabolite databases resulted in 468 metabolite measurements across studies. Some studies measure the same metabolite, leading to 260 unique metabolites across all studies. To be used in Mendelian randomization (MR), an exposure (a substrate in a reaction) needs to have at least one QTL available, resulting in 327 (206 unique) metabolites with at least one SNP (P *≤* 5 ⋅ 10^−8^). This is not a requirement when the metabolite is the outcome (the product in a reaction). An example of the MR result for the reaction between leucine and 4-methyl-2-oxopentanoate (supported by three databases). Leucine has genetic associations in 3 out of 4 mQTL studies where it was measured. We use SNPs in the associated regions for leucine as instruments to estimate the causal effect of leucine on 4-methyl-2-oxopentanoate. For brevity, causal estimates are only shown when the outcome is measured in Shin *et al*. Similar results are also found when the outcome is measured in Chen *et al*. All regional causal estimates (round circles) can be meta-analyzed into a weighted estimate (large diamond) for a joint causal estimate. **(c)**The ground truth positive causal relationships between metabolites extracted from 3 databases, containing 287 reactions across 154 metabolites. Causal estimates outside the pathway definitions are not shown. The size of the metabolite nodes represents the number of studies where the metabolite was measured. The width of the arrows represents how often a reaction was found in the three metabolic pathway definitions. The color denotes if a reaction was found or not. Green: The reaction was Bonferroni significant (*P* < 9.9 ⋅ 10^−7^) for MR-link-2 in at least one study combination when meta analyzing the estimates across the reaction (the weighted estimate from panel b). Grey: The reaction was not Bonferroni significant for MR-link-2. Pink: The substrate in the reaction does not have associated regions, meaning that there is no data for causal estimation.

Before setting out to understand the discriminative performance of MR methods on these metabolite networks, we performed a bias analysis and tested the *le Chatelier’s principle*. The bias analysis includes an additional 179 metabolite measurements that contain multiple measurements of 85 unique metabolites. We perform pairwise causal inference between measurements of the same metabolites in different studies (**Data S4**). Here, the expectation is that the causal estimate of a metabolite on itself is exactly 1.0 and any deviation from this value is considered as bias (**Fig. 4a-d**). We find that MR-link-2 has the smallest deviation from the expectation (*α*: 1.00) and thus the lowest estimation bias 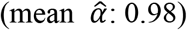 (**Fig. 4a**) compared to MR-IVW 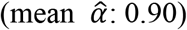 (**Fig. 4b**), MR-IVW LD 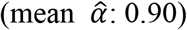 (**Fig. 4c**) and MR-PCA 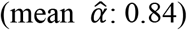 (**Fig. 4d**) (*23, 25, 34*) (**Data S5**) (**Table 1**). The chemical reactions present in human metabolism are governed by some well-established rules, one of which is the principle of *le Chatelier*, stating that an increase in a substrate will increase the product of a reaction (**Methods**) (**Supplementary Text**). Therefore, we expect that a causal estimate that represents a metabolic reaction should be strictly positive, as the causal effect represents the effect of the increase in a substrate. Indeed, when considering Bonferroni significant MR estimates (218,163 testable exposure, outcome and associated region combinations, *P* < 2.3 ⋅ 10^−7^), all methods identify more positive effects than negative effects (range: 59%-80%) (**Fig. 4e-h**), with the highest percentage (80%) for MR-link-2 (**Fig. 4e**), considerably higher than the second-best performing method, PCA-MR (63%) (**Fig. 4h**). Together these analyses indicate that the significant MR-link-2 estimates represent metabolism better than the significant estimates of other *cis* MR methods (**Fig. 4e-h**) (**Data S6**) (**Supplementary Text**).

**Fig. 4.**
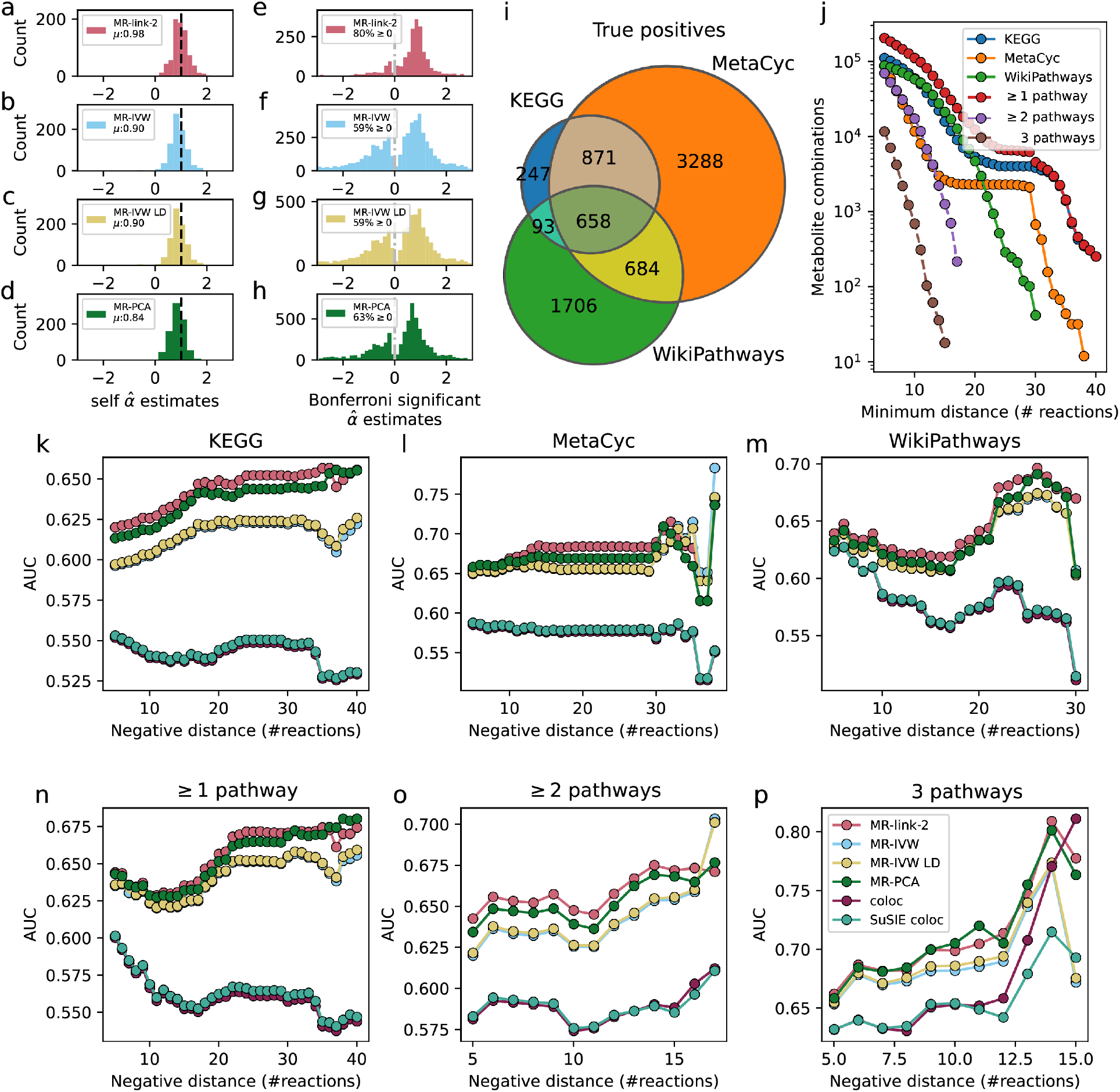
Comparison of different c*is* MR methods through effect size analysis, the true and false causal link datasets used for a comparison of discriminative ability of the metabolites in this study. Causal effects are estimated for an exposure based on each associated genomic region separately to test how reliable inferences are when only a single associated region is available. (**a-d**) The causal effect estimates of the Mendelian randomization (MR) methods tested in this study, when comparing nominally significant (*P ≤* 0.05) estimates between a metabolite on itself using two different mQTL datasets. The mean (μ) of a self-estimate is expected to be 1.0. Each panel is a different method: (**a**) MR-link-2 (402 comparisons), (**b**) MR-IVW (403 comparisons), (**c**) MR-IVW LD (404 comparisons) and (**d**) MR-PCA (412 comparisons). (**e**-**h**): The distribution of Bonferroni significant (*P ≤* 2.31 ⋅ 10^−7^) regional causal effect estimates for each method. We report the percentage of estimates that have a positive effect size, as positive effects are expected to represent direct metabolic reactions. When a substrate is physically converted into a product in a reaction, the causal effect should be strictly positive. (**a**) MR-link-2 (731 combinations), (**b**) MR-IVW (1753 combinations), (**c**) MR-IVW (1838 combinations) and (**d**) MR-PCA (1902 combinations). (**i**) A Venn diagram representing the number of true causal link combinations used for the regional results in this study. Separated by pathway definition. Each associated region for a substrate reaction is a positive for the regional results in this study. When two metabolites are more than one reaction away in the pathway databases, it is not considered as a true causal link. (**j**) The number of negatives (N) used in this study. As it is easy to confuse untested reactions with truly no reactions, we define the link between two metabolites as a negative (null) when they are separated by at least *m* reactions in the full metabolite graphs created from the databases (reaction distance). Only showing combinations when there are more than 10 negative links available. (**k-p**) The area under the receiver operator characteristic curve (AUC) of MR and colocalization methods benchmarked against different databases (**k-m**) and database combinations (**n-p**). Only showing comparisons when there are more than 10 negatives (same as panel **j**) per positive definition (same as panel **i**). When there is no SuSIE coloc estimate available for a region, we fall back to the original coloc estimate. True causal links and false causal links (**k**) from the KEGG pathway, (**l**) from the MetaCyc pathway, (**m**) from the WikiPathways pathway, (**n**) present in any pathway definition, (**o**) present in at least two pathway definitions, (**p**) shared in all pathways.

To compare the *cis* performance of all the methods in our metabolite network we assess each exposure locus independently, i.e., we do not meta-analyze any loci together as in **Fig. 3b**. This allows us to make per locus comparisons of coloc and different MR methods, which would be less transparent when meta-analyzed. True causal links are defined as direct reactions (**Fig. 3c**) (**Fig. 4i**). As it can be difficult to prove a negative in (human) biology, we utilize a reaction distance metric to define variable sets of false causal links that are increasingly likely to be null (**Fig. 4j**) (**Methods**). Compared to naively using all available non-causal combinations as false causal links, this approach reduces bias that may be due to causal relationships that exist between understudied metabolites, while also providing multiple AUC measures across different sets of false causal links. As such, this can be viewed as a sensitivity analysis due to imperfect definitions of a true null link dataset (**Fig. 4j**) (**Methods**). We ensure that the true null set is not too close to true links, by defining true null edges as those with the shortest path being at least five reaction long, while the strictest definition of null edges is defined as the maximum distance in which the number of false causal links is larger than ten (**Fig. 4j**) (**Methods**). If we compare the discriminative performance of the MR methods with coloc methods, we find that in aggregate, coloc methods have lower AUC than any MR method used here (**Fig. 4k-p**) (**Table 1**). Of note, generally the discriminative performance of coloc-SuSIE is better in our simulations, however, to ensure that methods use roughly the same amount of data, we fall back to the original coloc method when coloc SuSiE does not identify multiple causal variants (*14, 15*). This results in similar discriminative performance across comparisons (Pearson r: 0.969). The AUC of MR-IVW and MR-IVW LD are also very correlated (Pearson r: 0.992), as the LD corrected method produces identical results when there is only a single IV detected. Two methods use the whole genetic region for their inference, MR-PCA and MR-link-2, regardless of significance of the other genetic variants in the associated region. This approach is beneficial as MR-PCA or MR-link-2 usually (148 out of 156 AUC comparisons) provide the highest discriminative performance of all methods tested, with MR-link-2 being usually slightly better than MR-PCA: MR-link-2 has the highest AUC in 33 out of 36 comparisons in KEGG (**Fig. 4k**), 23 out of 33 in MetaCyc (**Fig. 4l**) and 25 out of 26 in WikiPathways (**Fig. 4m**) (**Data S7**) (**Methods**).

To reduce stochasticity caused by differences in pathway definitions, we also combined pathway references together (**Methods**) (**Fig. 4n-p**). MR-link-2 remains the method which is most often discriminative when all true causal links and false causal links are combined into a union (29 out of 36) (**Fig. 3j**) (**Fig. 4n**), when a true causal link and a false causal link is present in at least 2 datasets (12 out of 13) (**Fig. 4j**) (**Fig. 4o**). In the smallest true causal link dataset, the intersection of all true causal links and false causal links, MR-link-2 and PCA-MR have the highest AUC in 5 out of 11 cases each (**Fig. 4j**) (**Fig. 4p**) (**Data S7**).

The discriminative performance of MR methods also allows us to assess the characteristics of these pathway reference datasets. Considered individually, the MetaCyc pathway has the highest median AUC (0.684, MR-link-2), followed by KEGG (0.648, MR-link-2) and WikiPathways (0.639, MR-link-2) (**Fig. 4d**). When combining these pathway references, the intersection of all pathways has the highest median AUC (0.705, MR-PCA). However, the intersection is also the most unstable with the highest standard deviation across minimum reaction distances (maximum for coloc: 0.061) (**Data S7**). Indeed, the variability of the AUC estimates generally increases with the minimum reaction distance used for the null edge definition, which is where most rank changes between AUCs of methods were found (**Fig. 4k-p**). Indicating that as the datasets of false causal links reduce in size, the stability of the discriminative ability estimate also decreases.

Interestingly, in the KEGG and MetaCyc pathway references, the discriminative ability of MR-methods initially increases as the shortest path length to define null edges increases, coming to a plateau (**Fig. 4k, l**), suggesting that MR methods may have some power to detect metabolites that are linked through multiple reactions. Generally, MR-link-2 has increased discriminative ability over the other methods tested in this study. This is based on the AUC metric which is based on a segmentation of all P values or other test statistics. Generally, investigators researching causal molecular traits only consider *cis* regional Bonferroni significant (P < 2.3 ⋅ 10^−7^) results.

Therefore, we determine the precision and recall of the MR methods tested at this Bonferroni significance. We find that MR-link-2 has markedly improved precision in all the pathway comparisons (competing methods have 51-98% of the relative precision of MR-link-2, across all pathways and all methods), with lower recall (between 51-78 % of the recall for MR-link-2, across all pathways) (**Fig. S1A**). This indicates that significant results of MR-link-2 have a lower number of false positives than competing *cis* MR methods (**Fig. S1A**) (**Data S8**).

Up to now, we analyzed single locus estimates in isolation; however, MR estimates can be meta-analyzed together using the inverse of their variance estimate as weights (not to be confused with IVW MR methods) (**Fig. 3b**) (**Fig. 3c**) (**Table 1**) (**Methods**). When meta-analyzing results, discriminative performance of MR-link-2 is less pronounced over the other causal methods tested in this manuscript (101 out of 146 comparisons MR-link-2 does not have superior AUC) (**Fig. S2**) (**Data S9**). Moving away from the true causal links and negative causal links in the pathway definitions and considering the broader Bonferroni significant (48,567 exposure-outcome combinations, P < 1.0 ⋅ 10^−6^) results of these estimates weighted across associated regions, the precision remains highest for MR-link-2 in 5 out of 6 pathway comparisons. Unlike in the regional estimates, the recall of MR-link-2 is similar or higher than competing MR methods: in 2 out of 6 comparisons, MR-link-2 has superior recall and otherwise MR-link-2 has a recall that is 85-97% of the highest competing method (**Fig. S1B**) (**Data S8**). When comparing the 824 Bonferroni significant MR-link-2 results weighted across regions to the other tested methods, we find that MR-link-2 shows the least concordant results compared to other methods, with the lowest Jaccard index (min: 0.36, max: 0.47, lowest other Jaccard index: 0.66 between MR-PCA and MR-IVW) in a pairwise comparison of all four MR methods tested. These results suggest that MR-link-2 identifies causal relationships in a complementary manner compared to competing methods (**Fig. S3**), while retaining high precision and high recall. Indeed, MR-link-2 identifies 187 causal relationships at Bonferroni significance that are not found by other methods. This is more than for any competing method: 12 for MR-IVW, 4 for MR-IVW LD and 96 for MR-PCA. Fourteen of these 187 unique MR-link-2 estimates are found in the pathway databases used in this study whereas this number is only 4 out of 96 for MR-PCA, 1 out of 12 in MR-IVW and none of the four links unique to MR-IVW LD (**Data S10**).

### Biological interpretation of causal relationships between metabolites

We further explored the 187 Bonferroni significant MR-link-2 estimates (weighted across regions) between metabolites that are not reported by any of the three pathway references. (**Data S10**). Even though they may not necessarily be direct chemical reactions, causal relationships that MR-link-2 uniquely identified can be integrated into human metabolism, even if they do not represent a single reaction. One striking example is the negative bidirectional causal relationship identified between lactate and acetoacetate 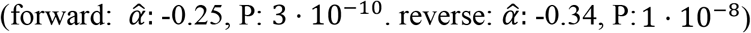. MR-link-2 is the only method that identified the negative causal relationship between the anaerobic fermentation pathway represented by lactate concentrations and the aerobic respiration pathway, represented by acetoacetate concentrations (*35, 36*) (**Data S10**). MR-link-2 is also the only method that identifies the positive causal relationship between two unsaturated fatty acids, (1-(1-enyl-stearoyl)-2-arachidonoyl-gpe (p-18:0/20:4) on cholesterol: 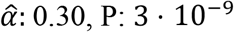 and n-stearoyl-sphingosine (d18:1/18:0) on cholesterol 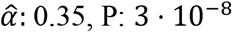). These two causal relationships recapitulate the role cholesterol has in stiffening membranes that contain unsaturated fatty acids (*37, 38*) (**Data S10**).

Another example is the regulation that MR-link-2 uniquely identified between s-1-pyrroline-5-carboxylate (P5C) and multiple amino acids. MR-link-2 identifies a causal relationship between P5C and arginine 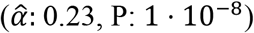 (**Data S10**). Interestingly, P5C is an intermediary in the biosynthesis of glutamate, arginine and ornithine, the latter two of which are integral parts of the urea cycle (*39*). Furthermore, MR-link-2 identifies a causal link between glycine and threonine as exposures and P5C as an outcome (glycine on P5C: 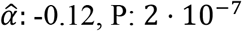, threonine on P5C: 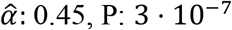) (**Data S10**). Here, MR-link-2 corroborates recent evidence that implicates glycine and P5C in a feedback mechanism, which if broken, causes hypomyelinating leukodystrophy-10 (HLD10) (MIM: 616420) (*40*). Patients suffering from HLD10 have a defective pyrroline-5-carboxylate reductase 2 (*PYCR2*) gene, which codes for the enzyme that converts P5C into proline. PYCR2 is an inhibitor of the serine hydroxy-methyltransferase 2 (SHMT2) enzyme that converts serine to glycine. Escande-Beillard *et al*. have demonstrated that HLD10 patients and *Pycr2* deficient mice have increased glycine in the brain, which can be rescued in mice through knockdown of the *Shmt2* gene, blocking the glycine buildup. MR-link-2 corroborates this evidence by showing a negative causal relationship between glycine and P5C. We hypothesize that this causal relationship functions through a link between glycine concentrations and PYCR2 activity. Where an increase in glycine also activates PYCR2, resulting in reduced P5C concentrations as it is converted into proline by PYCR2 (**Fig. S4**). Additionally, we show that there is a positive causal relationship between threonine and P5C. To the best of our knowledge, this relationship has not been investigated, and may further aid in the pathophysiological understanding the disease (*40*–*42*).

### Applying *cis* MR methods to complex traits

To ensure that MR-link-2 is not only effective at identifying causal relationships between metabolites, we tested the *cis* MR performance in a set of true and null relationships between complex traits. For this we offset complex trait combinations that are ‘considered causal’ with those that are considered ‘implausible or unsupported’ and ‘considered non-causal’ as defined by Morrison *et al*. (*9*) (**Methods**).

After applying *cis* MR methods to five trait combinations that are unlikely to be causal, e.g., outcomes that precede the risk factor in time, such as adult LDL-C levels impacting childhood onset asthma (COA). Only one false positive link is identified at nominal significance (by MR-PCA, LDL-C on COA, P: 0.02) (**Data S11**) (**Methods**). Perhaps as expected, all methods falsely identify the ‘non-causal’ relationship between HDL-C and coronary artery disease (CAD) as well as between HDL-C and stroke, which is notoriously difficult to accurately estimate through univariable MR methods (*9*) (**Fig. 5a**). When determining the per locus detection rate at nominal significance (P < 0.05), MR-link-2 has a consistently lower median T1E rate: MR-link-2: 0.089, MR-IVW: 0.151, MR-IVW LD: 0.168, MR-PCA: 0.157 (**Data S12**) (**Fig. 5a**). The lower T1E rate could be interpreted as lower power and indeed the detection rate is lower when analyzing causal relationships that are ‘considered causal’ (MR-link-2 median detection rate per locus: 0.207, MR-IVW: 0.267, MR-IVW LD: 0.267, MR-PCA: 0.251) (**Fig. 5b**). These results show that the relative loss in detection power for MR-link-2 compared to other methods is considerably lower than the decrease in T1E rate. We decided to investigate how *cis* methods behave in the HDL-C to CAD analysis, which all *cis* MR methods identify as significant. It is widely accepted that the shared genetics of LDL-C and HDL-C cause MR to identify a causal relationship between HDL-C and CAD, while this is due to pleiotropy from the true causal link of apolipoprotein B and CAD (*6*). This conclusion came from the landmark Voight *et al*. paper, that analyzed this relationship in a protein coding locus of the *LIPG* gene that was exclusively associated to HDL-C. With larger sample sizes, this locus is now associated to LDL-C and non-HDL-C (*43*). Even when only using SNPs from the *LIPG* locus as instruments, all methods spuriously find a causal effect of HDL-C on CAD (max P: 0.007, MR-IVW). Inspired by the Voight *et al*. analysis, we isolated HDL-C associated regions that are not associated to other cholesterol traits (*6*). Interestingly, all methods tested remain significant in meta-analysis. When adding a minimum distance of an HDL-C region to the other cholesterol regions, MR-link-2 requires the shortest distance between regions for the meta-analysis to reach a non-significant P value (> 0.05) (**Fig. 5c**). Interestingly, there is no initial sign of loss-of-power in this analysis, as MR-link-2 initially starts off with more significant test statistics than MR-IVW and MR-IVW LD (**Fig. 5d**) (**Data S13**). Upon the meta-analysis of all loci for all complex trait combinations analyzed in this study, we find that MR-link-2 has substantially lower heterogeneity in terms of Cochran’s Q statistic (median for MR-link-2: 586, lowest competing: 1359) (**Methods**) (**Fig. 5d**) (**Data S11**). The low heterogeneity statistic of meta-analysis could be attributed to MR-link-2’s low false positive rates, which have more realistic standard error estimates, possibly due to accounting for pleiotropic effects.

**Fig. 5.**
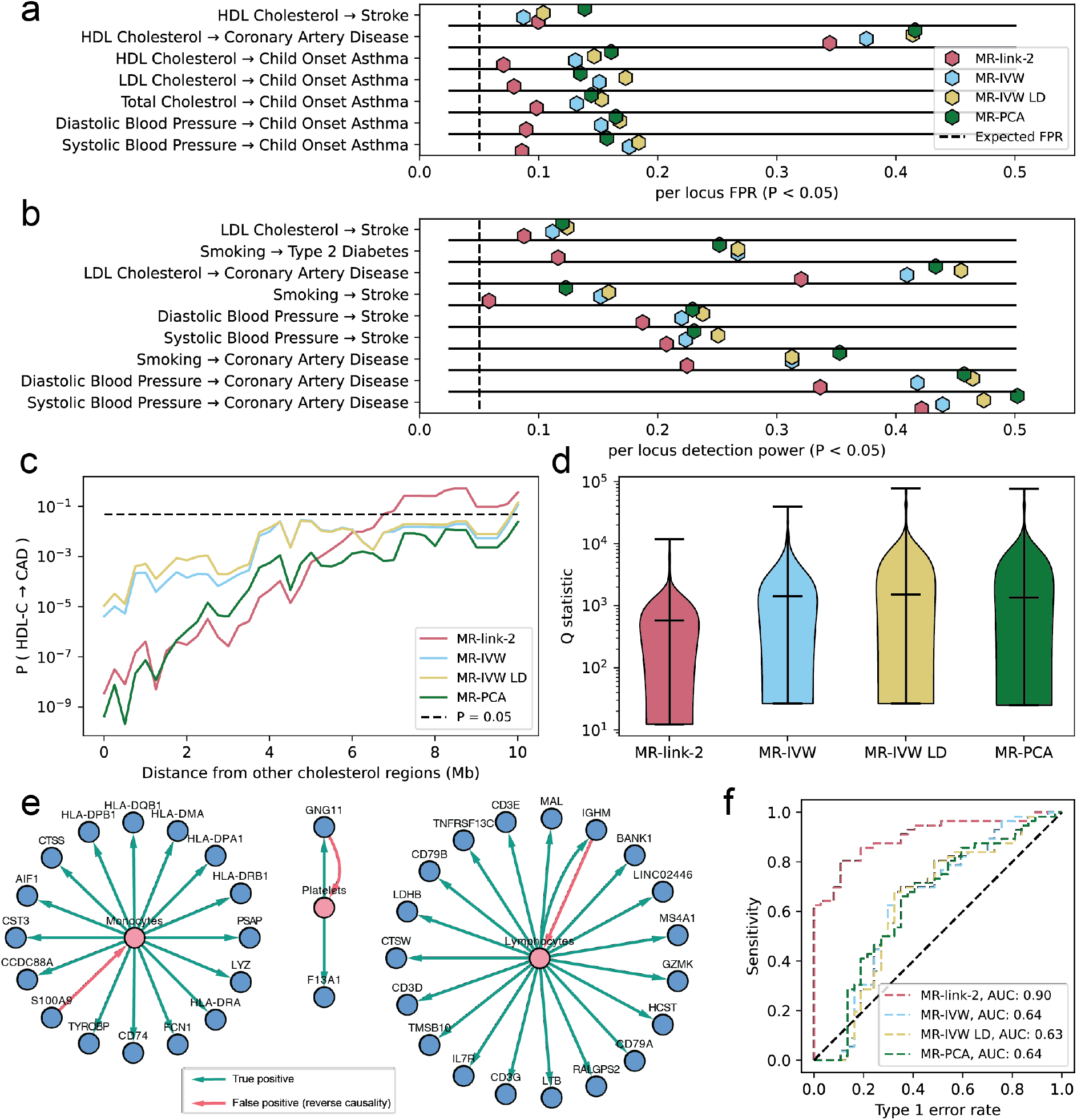
Analysis of different MR methods on canonical causality and the causality between blood cell traits. (**a**) The per locus detection rate (at P < 0.05) for phenotype combinations that are not considered causal or are unlikely to be considered causal by Morrison *et al*. (**b**) The per locus detection rate (at P > 0.05) for phenotypes that are considered causal. (**c**) P value (log10 scale) for a meta-analysis between HDL-C and coronary artery disease when the HDL-C loci are a minimum distance from other cholesterol loci (y-axis). (**d**) Violin plot of the heterogeneity statistics of the MR methods tested in this study. Upon meta-analysis of all the phenotype combinations in (a) and (b), we plot the Q statistic for each method (log10 scale). (**e**) Blood cell type and eQTL analysis results. MR-link-2 Bonferroni significant (P < 5.15 ⋅ 10^−4^) causal links between cell type concentrations and the RNA expression of their respective marker genes (**Data S17**). Green colored arrows indicate the cell type influences the RNA gene expression in blood causally. These are considered true causal links. The red arrows indicate an (incorrect) causal link between the gene expression and the blood cell type marker, indicating reverse causality. (**f**) Area under the receiver operator characteristic curve for the cell type directionality analysis for all MR methods tested in this study based on the reported P value of the method.

### Using reverse causality as true causal links from eQTL information

To ensure that MR-link-2 does not identify reverse causality excessively, we applied the studied *cis* MR methods to test how often we can identify the correct causal direction between whole blood bulk gene expression levels and blood cell composition phenotypes (*44*–*46*) (**Fig. 1f**). Our assumption is that when analyzing the mixture of cells such as blood, the differences in the concentrations of certain cell type causally affect the gene expression of respective marker genes (i.e., the observed expression quantitative trait locus (eQTL) effect is caused by genetically regulated cell type differences), whereas the opposite direction is a false positive. These marker genes are derived from reference single cell RNA expression experiments that are typically used to identify cell types from untargeted assays (*47*).We test the bidirectional causal relationship between RNA expression of 73 cell-type marker genes and 12 types of peripheral blood mononuclear cells (PBMCs) measured in up to 563,085 individuals of European ancestry (*47*). Using newly generated RNA expression eQTLs from the eQTLGen consortium that contains a genome-wide *cis* and *trans* eQTL summary statistics from 19 cohorts and 14,855 individuals (*48*).

Upon the meta-analysis of MR estimates across associated regions for 93 true (cell type to gene expression) and 93 false (gene expression to cell type) causal combinations, we find 39 Bonferroni significant (*P* < 5.3 ⋅ 10^−4^) MR-link-2 comparisons. All estimates have a positive causal effect direction estimate (**Data S14**). Indeed, we find that 94.9% (all but three) of the Bonferroni significant MR-link-2 estimates are in the correct direction (from cell type to RNA expression) (**Fig. 5e**). Interestingly, in these Bonferroni significant estimates, two false positives are bidirectional, meaning that MR-link-2 identifies both the true causal link and the false causal link (**Fig. 5e**). In comparison with the other tested *cis* MR methods, we find that MR-link-2 has much higher discriminative ability (MR-link-2 AUC: 0.90, best competing: 0.64) to identify the correct effect direction based on the MR methods causal estimate P value (**Fig. 5f**) (**Methods**). This may not be surprising, as the reverse causal analysis is a special case of pleiotropy, which the other tested MR methods do not explicitly account for.

We were intrigued by the reverse causal direction that MR-link-2 identified between *S100A9* expression and monocyte concentrations. *S100A9* is considered a marker gene for monocyte concentrations based on single cell experiments (*47*). Interestingly, MR-link-2 only identifies the reverse effect: *S100A9* increases monocyte concentration with a larger effect (*S100A9* as exposure: 1 locus, 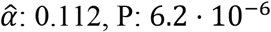, monocyte count as exposure 514 loci, 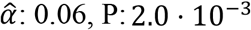 (**Fig. 5f**) (**Data S14**). Based on a literature review, the causal effect direction of *S100A9* may actually have been correctly estimated by MR-link-2, as *S100A9* has been shown to promote accumulation of leukocytes from mouse knockout experiments (*49*) as well as inhibiting dendritic cell differentiation (*50*). Dendritic cells are a class of monocytes, which upon differentiation, migrate to non-blood tissues, reducing monocyte blood concentrations. These results suggest that *S100A9* expression promotes monocyte accumulation in whole blood and might represent a true causal link, as found by MR-link-2.

## Discussion

In this study, we present a new *cis* MR method: MR-link-2, which can perform pleiotropy robust MR in a single region when summary statistics and an LD reference are available. To our knowledge, MR-link-2 is the only summary statistics *cis* MR method that explicitly models pleiotropy. MR-link-2 provides estimates of causal relationships through modeling the effect of all SNPs in an entire *cis* region on an exposure and an outcome. We have tested MR-link-2 in four different validation datasets, three of which being based on real data. When compared to competing *cis-*MR and colocalization methods, MR-link-2 has better discriminative ability and lower T1E. Furthermore, MR-link-2 uniquely identified compelling biological examples, such as the negative causal relationship between lactate and acetoacetate which contrasts anaerobic and aerobic energy pathways in humans, respectively, or the negative relationship between P5C and glycine, which has been experimentally found in model systems to be key in the HLD10 syndrome. Together, these results illustrate the ability of MR-link-2 to help shape our understanding of the underpinnings of molecular mechanisms in humans, including those underlying disease.

Our simulations suggested that MR-link-2 is sensitive to scenarios with strong LD between causal SNPs, especially when the reference LD matrix is measured with high uncertainty, in which case other *cis* genetic methods outperformed MR-link-2. However, in real data validations MR-link-2 exhibits lower T1E rates compared to other methods, indicating that the most extreme scenarios of our simulations are unlikely to occur in human biology. The application to real-world validation datasets show that MR-link-2 is robust to the presence of horizontal pleiotropy in a locus. We make this conclusion based on the lower false positive rates in the per locus estimates of complex traits, the closer agreement with *le Chatelliers* principle in metabolites, the lower heterogeneity of estimates in our complex trait application and the identification of causality in the correct direction in the blood cell count and eQTL analysis. However, this does not mean that MR-link-2 is robust to all violations of the assumptions underlying MR. In cases of extreme pleiotropy, e.g., the HDL-C to CAD analysis, T1E rates are increased. In less extreme cases, we expect the per locus T1E to be 0.05, while our median per locus T1E is 0.09 in the non-causal complex trait analysis. This analysis indicates that accounting for pleiotropy could be improved by allowing for multiple exposures in the model, which could be a natural extension of MR-link-2. Taken together and given that MR-link-2 i) provides meaningfully different results from the other tested methods in this study and ii) has lower T1E than other methods, we believe that MR-link-2 can be useful both as a standalone method as well as for secondary validation. Even though the MR methods tested in this work are developed to be principally for the analysis of *cis* associations, they still perform well when meta-analyzing multiple *cis* estimates together. We have found no limitation to the number of regions that MR-link-2 estimates, enabling its application to complex exposures.

The true and null causal relationships from the reference datasets used in this study can be discriminated by genetically informed causal inference methods. Unfortunately, these reference datasets remain imperfect, which is illustrated by the limited overlap between pathway references in the metabolite datasets. Another striking example relates to *S100A9*, for which we hypothesize that the impact of changes in expression are causal to changes in blood composition. This is at odds with our initial assumption that changes in blood composition are causal to changes in the expression of cell marker genes. The benchmarking data developed in this study provide a reasonable gold-standard for testing causal inference methods. In the future, the community should increasingly seek to refine and broaden these datasets to facilitate the development and benchmarking of new causal inference tools. One important step towards this is to build causal datasets in a tissue specific way. The current ground truth datasets will contain some tissue specific links that will be difficult to detect. Indeed, the limited tissue specificity in the metabolite analysis may contribute to a the relatively low recall of all causal edges. Improvement of the tissue specificity in these datasets may contribute to better discriminative ability of each method.

In this study we use summary statistics of European ancestry individuals when they are available. We justify this decision based on sample sizes and availability of a large and representative LD reference. It is important to note that our methodology is not limited to individuals of European ancestry. We even hypothesize that a more diverse association panel would make MR-link-2 more powerful as the diversity of the available LD panel will allow for better distinction between causal effect and horizontal pleiotropy which we believe is paramount to correct causal inference. The availability of more multi-ancestry population studies is likely to improve our understanding of causality, with the caveat that the LD panel should match the ancestry composition from which the summary statistics were derived.

In all the validations performed in this study, MR-link-2 has shown to be a promising tool to study exposures with dominantly *cis*-associations and leads to improved identification of causal links between molecular trait, which will – in turn – facilitate shedding light on the molecular basis of complex traits.

## Supporting information

Table S6 in tab separated txt.gz format

Supplementary tables S1-S5 and S7-S16 combined in a zip format. The included files are in the xlsx file format.

## Data Availability

Data and materials availability: The code for simulations and working examples for MR- link-2 and the other cis- causal inference methods are available at: https://github.com/adriaan-vd-graaf/mrlink2. The summary statistics for the metabolite analysis, the complex trait analysis and the blood cell type composition phenotypes are available from the respective source publications (Data S15). The data availability of each of 15 the eQTLGen Consortium cohorts is listed in the (Supplementary Text). The genotype information underlying the LD matrices for the UK10K data resource were downloaded from (EGAD00001000740, EGAD00001000741).

## Acknowledgements

We would like to thank all the study participants for their altruistic donations of their biological materials. Acknowledgments of the cohorts of the eQTLGen Consortium, we refer to the **Supplementary Text**

## Author contributions

Conceptualization: ZK, AvdG

Methodology: ZK, AvdG, RW, UV, LF, CB, EC

Investigation: ZK, AvdG, RW, UV, LF

Visualization: AvdG

Funding acquisition: ZK, LF, CB

Project administration: ZK

Supervision: ZK

Writing – original draft: Avdg

Writing – review & editing: CA, CB, ZK, LH, RW, UV, AvdG

## Competing interests

Authors declare that they have no competing interests.

## Data and materials availability

The code for simulations and working examples for MR-link-2 and the other *cis-* causal inference methods are available at: https://github.com/adriaan-vd-graaf/mrlink2. The summary statistics for the metabolite analysis, the complex trait analysis and the blood cell type composition phenotypes are available from the respective source publications (**Data S15**). The data availability of the each of the eQTLGen Consortium cohorts is listed in the (**Supplementary Text**). The genotype information underlying the LD matrices for the UK10K data resource were downloaded from (EGAD00001000740, EGAD00001000741).

**Supplementary Materials**

Methods Supplementary

Text Figs. S1 to S4

Data S1 to S16

## Methods

### Mendelian randomization and colocalization methods

#### Existing cis genetic methods used in this study

We use six genetic methods in this study to identify if two phenotypes are causally related to each other. Two methods are colocalization methods and four methods are MR based methods.

We use two versions of colocalization, namely coloc (*14*) and coloc-SuSIE (*15*). Both methods test for the following question: Are the causal variants of the traits shared? coloc assumes that there is a single causal variant in the locus, whereas coloc-SuSIE relaxes this assumption by identifying independent SNPs and subsequently performing a conditional coloc analysis per variant. We define detection of a causal variant sharing as having a coloc posterior probability (PP4) larger than 0.9 for the 4^th^ hypothesis (two traits share a causal variant). As coloc-SUSIE may estimate multiple causal effects, we take the maximum PP4 across analyses (**Table 1**).

We use three existing (*cis*) MR methods and introduce one new MR method: MR-link-2. The three existing MR methods used are MR-IVW (*34*), MR-PCA (*23*) and MR-IVW LD (*23*). We used the ‘mr_ivw’ and ‘mr_wald_ratio’ functions (for multiple instrumental variables and a single instrumental variable respectively) from the TwoSampleMR package (*51, 52*) ‘https://github.com/mrcieu/TwoSampleMR‘. We adapted the MR-PCA and MR-IVW-LD code from Burgess and Thompson (*23*) our adaptation was limited to storing duplicated code segments in memory that can otherwise take a long while to process. (reference on https://github.com/adriaan-vd-graaf/mrlink2). We compared our adaptation to the original and found no difference in effect estimates or levels of significance (**Table 1**). MR-IVW-LD accepts the same variants as instrumental variables; however, the method adjusts their effect sizes based on the LD in between the instrumental variables.

When determining detection rates in our simulations, we consider it evidence for a causal relationship if the MR P value is smaller than 0.05. We have not compared the original MR-link (v1) method as our analysis depends exclusively on summary statistics and as such, the method is not suited for our comparisons (*20*).

#### The MR-link-2 likelihood function

MR-link-2 is a likelihood function that models the summary statistics found between a cohort of *n*_*X*_ exposure phenotypes (*X*) and an *n*_*Y*_ outcome phenotypes (*Y*). The full derivation of the MR-link-2 likelihood function can be found in the **Supplementary Text**. We continue with a bird’s eye view of the full derivation.

We model the causal relationship *α* between an exposure and outcome in the following way:

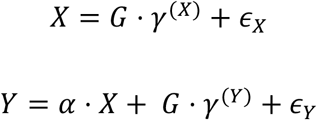

Here, *G* represents a genotype matrix, with normalized genotypes to zero-mean and unit variance across samples. SNP effects are modelled as random, 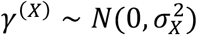, where 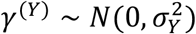 is related to the *cis* heritability of *X*, such that 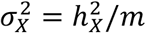 and 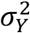 is the direct (vertically pleiotropic) *cis* heritability of *Y*, such that 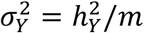. Error terms are distributed as follows: 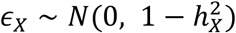 and 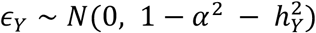. The marginal GWAS summary statistics, estimated in samples of size *n*_*X*_ *and n*_*Y*_, respectively, can then be written as:

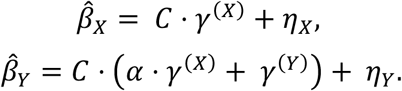

Where *C* represents an *m* by *m* LD matrix: *C* = *G*^*T*^ ⋅ *G /n* and *η*_*X*_ = *α* ⋅ (*G*^*T*^ ⋅ ϵ_*X*_)*/n*_*X*_ which is distributed as 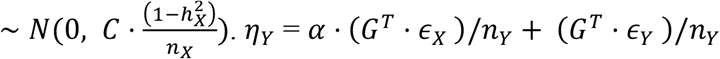 is distributed as 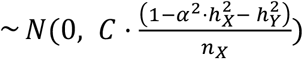. These distributional assumptions allow us to formulate a joint likelihood function for both summary statistics to estimate the causal effect and underlying multivariable SNP effects: 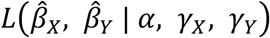. To maximize the likelihood function, we would have to optimize 2 ⋅ *m* + 1 variables which can be difficult to optimize, as *m* could contain thousands of parameters, when an associated region has many genetic variants. Therefore, we integrate out the underlying SNP effects *γ*^(*X*)^ and *γ*^(*Y*)^, conditional on the per SNP heritabilities 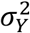 and 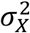. This reduces the parameters to optimize to 3, which is faster and increases power. After some algebraic transformations (**Supplementary Text**), the MR-link-2 log likelihood function simplifies to:

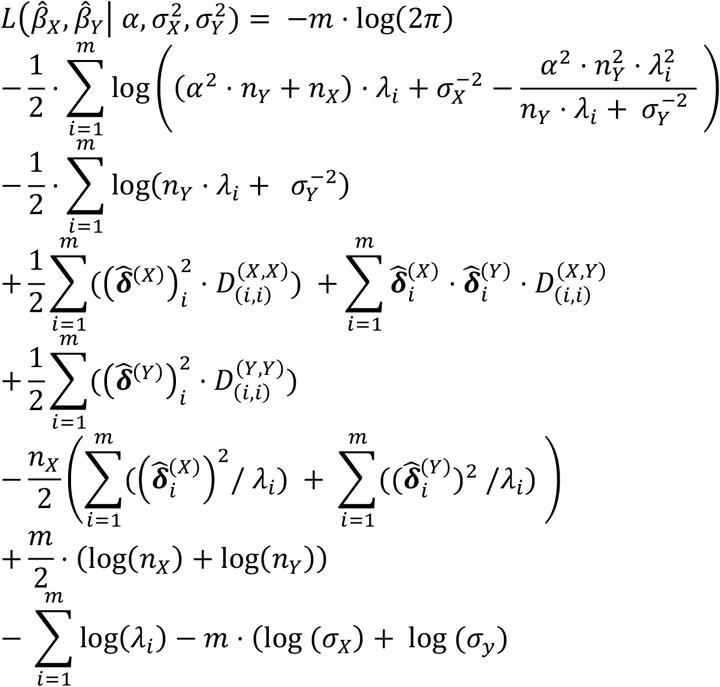

To arrive to this expression, we used a singular value decomposition of the correlation matrix,*C* = *U* ⋅ Λ ⋅ *U*^*T*^, which preserved 99% of the variance. This led to the introduction of the following quantities: *λ*_*i*_ is the *i*^th^diagonal element of Λ, 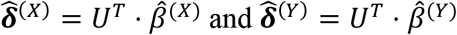. Finally, the *D*^(*X*,*X*)^, *D*^(*X*,*Y*)^ and *D*^(*Y*,*Y*)^ are diagonal matrices with diagonal elements defined as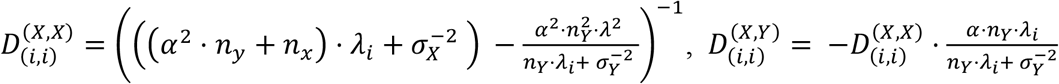 and 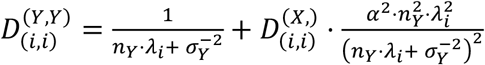.

#### Application of the MR-link-2 likelihood function

We optimize the MR-link-2 likelihood function using the Nelder-Mead optimizer using the `scipy optimize minimize` function (*53*). We optimize the likelihood function three times, first by setting i) *α* = 0 and freely estimating 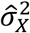 and 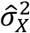. Then, ii) by setting the pleiotropic variance 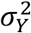 to zero and freely estimating 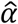 and 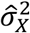. And finally, iii) by freely estimating all three parameters 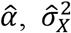 and 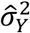. We identify confidence intervals and P values of 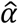, and 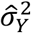 through a likelihood ratio test with one degree of freedom.

A full implementation for MR-link-2 is available online at https://github.com/adriaan-vd-graaf/mrlink2. This implementation accepts 2 harmonized summary statistic files and a plink style “.bed” genotype file used for generating an LD reference (*54*). For all associated regions in the exposure summary statistics file, MR-link-2 provides a causal estimate.

### Simulations

#### Simulations of summary statistics

We performed extensive simulations to ensure that MR-link-2 provides accurate causal inference, as well as to compare it to other *cis* methods. Our simulations were performed with the goal of mimicking a *cis* region of a molecular -omics study that is potentially causal to a complex trait that is measured in a large cohort. Therefore, the exposure is measured in 10,000 individuals (*n*_*X*_), while the outcome is measured in 300,000 individuals (*n*_*Y*_) in a genomic region of 2,068 SNPs (*m*) that is derived from a UK10K region on chromosome 10 (*22*). Our simulations contain six different parameters that we vary: the causal effect (*α* ∈ {0,0.5,0.1,0.2}), the exposure heritability 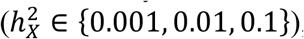, pleiotropy that is represented as outcome heritability 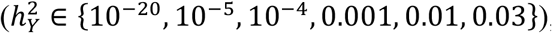, The size of the linkage disequilibrium (LD) reference (*n*_*ref*_ ∈ {500,5000,∞}), the number of underlying causal SNPs (*m*_*casual*_ ∈ {1,3,5,10,100}), the minimum and maximum LD between causal and pleiotropic SNPs 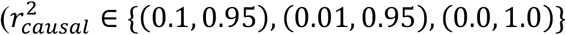 for minimum and maximum correlation respectively). In total we have simulated 2,700 different scenarios with 1,000 replications per scenario. Of note, none of the 2,700 parametrizations of our simulations do not violate the specific assumptions underlying MR-link-2. MR-link-2’s underlying assumptions are violated when *n*_*ref*_ ≠ ∞, *m*_*casual*_ ≠ 2,068 = *m* and when 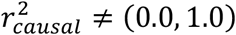.

We simulated summary statistics for the two phenotypes in the following way:

1. A total of *m*_*casual*_ SNPs are selected from the region for the exposure and the outcome. Selection is random across the region when *r*_*casual*_ = (0, 1) and following the procedure of the original MR-link manuscript otherwise (*20*). In this procedure, SNPs are selected iteratively until *m*_*casual*_ SNPs are selected. The first SNP is drawn randomly from the region, afterwards the next SNP is drawn from all possible SNPs that meet the correlation criteria compared to all other previously selected SNPs.
2. Independent SNP effects for the exposure and the outcome (*γ*_*X*_ and *γ*_*Y*_ respectively) are randomly drawn from a normal distribution 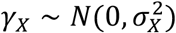 and 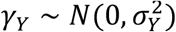 for each SNP that is selected to be causal, otherwise it is set to zero.
3. Independent SNP effects *γ*_*Y*_,*γ*_*X*_ are transformed into unconditional effect sizes *β* in the following way. We multiply the independent SNP effects by the correlation matrix *C* and add measurement error term 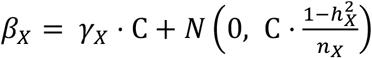and 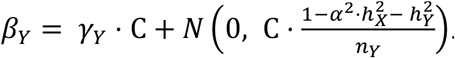.

These simulated summary statistics are then introduced in their respective MR and colocalization algorithms. When *n*_*ref*_ is infinite, the LD matrix that is used as input for the algorithms 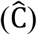 is the same as the original (*C*). When *n*_*ref*_ is not infinite, we simulate imprecisely measured LD through Wishart sampling the C matrix (*55*). In cases where Σ is not positive semidefinite, we add regularization constants (up to 0.5) to the diagonal of the original matrix to ensure that Wishart sampling continues correctly.

For the methods that require instrument selection (MR-IVW and MR-IVW LD) we selected instruments used P value clumping at a P value threshold of 5 ⋅ 10^−8^ and an LD r^2^ squared threshold of 0.01.

### Summary statistics used in this study

#### Summary statistics harmonization and associated region selection

In this work we utilize summary statistics from a variety of different studies. We processed summary statistics of all studies in the same way: First, if necessary, we lifted over summary statistics into human chromosome build 37 using UCSCs liftover tool (https://genome.ucsc.edu/cgi-bin/hgLiftOver) combined with their chain files (https://hgdownload.soe.ucsc.edu/downloads.html). Then, we include SNP variants that have LD information available, by overlapping the variants (based on chromosome, position and alleles) present in the summary statistics file with the variants in our LD reference (UK10k). Due to potential strand inconsistencies, palindromic SNPs were removed.

We ensured that effect size magnitudes of summary statistics are the same between studies by converting to standardized effect sizes:

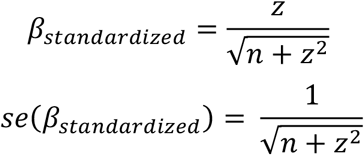

Where the *n* is the sample size of the tested SNP and *z* is the Z score of the tested SNP. If *n* was not available per SNP, we set *n* to be the maximum sample size reported by the authors. If *z* was not available, the P value of the SNP-trait association combined with the effect direction was converted into a Z score. Genetic associations are retained if they have at least a minor allele frequency of 0.5% in the UK10K LD reference and if the variant has been measured in at least 95% of the maximum number of measured individuals (if the information was available).

We identify associated regions using the --clump command of plink (v1.90b7) (*54*), using a clumping window of 250Kb, an LD threshold of 0.01 *r*^2^ and a P value threshold of 5 ⋅ 10^−8^ If clumped regions overlap, we combine them together, so these regions can be much larger than 250Kb. All harmonized regions are then analyzed by each *cis* genetic method individually. If the method requires the selection of IVs (MR-IVW, MR-IVW LD), these are clumped inside the region at a P value threshold of 5 ⋅ 10^−8^ and an r^2^ threshold of 0.01.

### Summary statistics of metabolite QTL studies

We analyzed the summary statistics of four different mQTL studies. To match the associations to our reference panel, we chose to analyze the European component of the individuals, when available. Shin *et al*. was downloaded from http://metabolomips.org/gwas/index.php?task=download (*33*), Lotta *et al*. was downloaded from https://omicscience.org/apps/crossplatform/ (*32*), Chen et al summary statistics derived from European populations were downloaded from the GWAS catalog (*31, 56*). The UK biobank summary statistics were downloaded from the IEU open GWAS project, where we included 19 accessions that represented small metabolites (*30, 57*). More information about the metabolites used in this study can be found in (**Data S4**).

### Summary statistics of complex trait harmonization and processing

To understand the behavior of *cis* MR methods, we selected ground truth (non-)causal relationships between complex trait combinations from Morrison *et al*. (*9*) These involve 10 unique phenotypes, summary statistics of which were downloaded from their respective datasets (**Data S15**). In brief, we utilized summary statistics from lipid phenotypes (LDL-C, HDL-C and total cholesterol) from Graham *et al*. (*43*), blood pressure (diastolic blood pressure and systolic blood pressure) summary statistics from Warren *et al*. (*58*), coronary artery disease summary statistics from Aragam et al. (*59*), stroke summary statistics from Mishra *et al*. (*60*), childhood asthma summary statistics from Ferreira *et al*. (*61*), Type 2 diabetes summary statistics from Mahajan *et al*. (*62*) and summary statistics of smoking from Karlsson Linnér *et al*. (*63*). Some of these studies are based on multi-ancestry analyses, when population specific summary statistics were available, we exclusively analyzed the European subset of the final summary statistics (**Data S16**).

### Summary statistics of eQTLGen

The eQTLGen Consortium is an initiative to investigate the genetic architecture of blood gene expression and to understand the genetic basis of complex traits. We used interim summary statistics from eQTLGen phase 2, wherein a genome-wide eQTL analysis has been performed in 19 cohorts, representing 14,855 individuals.

All 19 cohorts performed cohort-specific analyses as outlined in the eQTLGen analysis cookbook (https://eqtlgen.github.io/eqtlgen-web-site/eQTLGen-p2-cookbook.html). Genotype quality control was performed according to standard bioinformatics practices and included quality metric-based variant and sample filtering, removing related samples, ethnic outliers and population outliers. Genotype data was converted to genome build hg38 if not done so already and the autosomes were imputed using the 1000G 30x WGS reference panel (*64*) (all ancestries) using our imputation pipeline (https://github.com/eQTLGen/eQTLGenImpute). Like the genotype data, gene expression data was processed using our data QC pipeline (https://github.com/eQTLGen/DataQC). For array-based datasets, we used the results from empirical probe mapping approach from our previous study (*21*) to connect the most suitable probe to each gene which has previously been to show expression in the combined BIOS whole blood expression dataset. Raw expression data was further normalized in accordance with the expression platform used (quantile normalization for Illumina expression arrays and TMM (*65*) for RNA-seq) and inverse normal transformation was performed. Gene expression outlier samples were removed and gene summary information was collected for filtering at the central site. Samples for whom there were mismatches in genetically inferred sex, reported sex, or the expression of genes encoded from sex chromosomes were removed. Similarly, samples with unclear sex, based on genetics or gene expression were removed.

The HASE framework (*66*) was used to perform genome-wide meta-analysis. For genome-wide eQTLs analysis, this limits the data transfer size while ensuring participant privacy. At each of the cohorts, the quality controlled and imputed data was processed and encoded so that the individual level data can no longer be extracted, but while still allowing effect sizes to be calculated for the linear relationship between variants and genes. (https://github.com/eQTLGen/ConvertVcf2Hdf5 and https://github.com/eQTLGen/PerCohortDataPreparations).

Centrally, the meta-analysis pipeline was run on the 19 cohorts. The pipeline which performs per cohort calculations of effect sizes and standard errors and the inverse variance meta-analysis is available at https://github.com/eQTLGen/MetaAnalysis. We included 4 genetic principal components as covariates. Per every dataset, genes were included if the fraction of unique expression values was equal or greater than 0.8, Variants were included based on imputation quality, Hardy-Weinberg equilibrium and minor allele frequency (MAF) (Mach R^2^≥0.4, Hardy-Weinberg P≥1×10^−6^ and MAF≥0.01).

### Summary statistics of cell type proportions

We processed the summary statistics of 15 cell type composition phenotypes from the Chen et al. (2020) meta-analysis, using the. Summary statistics of individuals with European ancestry. these cell type composition phenotypes were downloaded from http://www.mhi-humangenetics.org/en/resources/ (*44, 45*).

### Metabolite analysis

#### Harmonization of metabolites

The mQTL studies studied here use different metabolite naming schemes for their metabolites. To make sure that all metabolites studied are the same, we harmonized metabolites to HMDB database identifiers (*67*). The HMDB is a large reference database for human metabolites and contains references to other metabolites. In this work we utilized the HMDB database of 17^th^ of November 2022. Downloaded from https://hmdb.ca/system/downloads/current/hmdb_metabolites.zip.

As a starting point for the harmonization of metabolites, we utilized the metabolite comparison information provided by the supplementary table 4 of the Chen *et al*. publication (*31*). Here, 2,075 metabolites across 6 studies were provided a harmonized name. One mQTL study used in this work was not considered (UKB metabolites), therefore, we manually harmonized 19 metabolites from the unharmonized UKB information into a derived table (**Data S4**). Combined, the 4 mQTL studies under investigation have 1,518 unique harmonized metabolites measured.

We removed 430 measured metabolites that were not matchable to a single compound, leaving 1,037 metabolites for study. 660 / 1,037 metabolites were already matched to the HMDB database by Chen *et al*. For the remaining matches, we matched name and synonyms for a further 239 matched names to HMDB identifiers. As a final step we manually matched a further 56 compounds based on manual web searching the HMDB website, resulting in 854 metabolites with a HMDB identifier (**Data S4**). To ensure that metabolites can be easily matched with other databases, these metabolites have also been matched with InChIKey, KEGG compound ID and ChEBI ID (**Data S4**).

#### Metabolite networks used in this study

We created 3 different metabolite networks to benchmark our causal inference method: the MetaCyc HumanCyc v24 pathway (released 30th of April 2020), the WikiPathways Homo sapiens (“https://wikipathways-data.wmcloud.org/current/gpml/wikipathways-20230510-gpml-Homo_sapiens.zip”) pathway and the KEGG pathway (downloaded on the 30^th^ of May 2023). Each pathway has an *extended graph* and a *measured graph*. The measured graph contains the direct reactions of the measured metabolites in this study, whereas the extended graph contains all the causal relationships in the pathway definition.

#### Creation of the MetaCyc network graph

To create the MetaCyc metabolite network we loaded the compound and reaction information from downloaded MetaCyc HumanCyc flat files (*27*). Some compounds are very common reactants, therefore, we removed the following HumanCyc Identifiers from our analysis: {‘GTP’, ‘CL-’, ‘CYS’, ‘Fatty-Acids’, ‘HCO3’, ‘GDP’, ‘3-5-ADP’, ‘MALONYL-COA’,’NADH-P-OR-NOP’, ‘CMP’, ‘PAPS’, ‘NAD-P-OR-NOP’, ‘SUC’, ‘Acceptor’, ‘AMMONIUM’, ‘NA+’, ‘ACETYL-COA’, ‘ADENOSYL-HOMO-CYS’, ‘HYDROGEN-PEROXIDE’,’UDP’, ‘AMP’, ‘Donor-H2’,’NADH’, ‘PPI’, ‘NADPH’, ‘ADP’, ‘NAD’, ‘CARBON-DIOXIDE’, ‘Pi’, ‘CO-A’, ‘NADP’, ‘ATP’,’OXYGEN-MOLECULE’, ‘WATER’, ‘PROTON’}. After common reactant removal, we built an extended graph containing 2,560 compounds and 3,900 reactions. We matched HumanCyc compound identifiers with HMDB and ChEBI identifiers (*27, 68*). After matching with our mQTL studies, this resulted in 115 compounds across 146 reactions in the measured graph.

#### Creation of the WikiPathways network graph

To create the WikiPathways metabolite network, we downloaded each individual human pathway and kept all combinations where a compound is converted into another according to the ‘mim-conversion’ arrow specifier (*29*). This resulted in an extended graph containing 3,795 compounds and 4,871 reactions. We matched the compounds in WikiPathways with the HMDB (*67*) or the ChEBI databases (*68*) and find 160 compounds across 155 reactions that were measured in the measured graph.

#### Creation of the KEGG network graph

To identify the KEGG metabolite network, we used the following procedure. For the 435 compounds for which a KEGG ID was matched, we downloaded the compound data and determined in which full pathways (‘map’) the compound could be found. For each of these 229 pathways, we downloaded the human equivalent (replacing ‘map’ with ‘hsa’) KGML files. From these KGML files, all human reactions were parsed to construct a graph of 1,877 reactions across 1,270 compounds in the extended graph. 113 of which were measured in at least one mQTL study. This measured graph contains 126 reactions.

#### Bias estimation and le Chatelliers principle in mQTL MR

We test the bias of MR methods by comparing causal estimates of the same metabolite when they are measured in different studies (**Fig. 4a-d**). We take the mean of the effect sizes of these ‘self comparisons’ when the respective MR method identifies the causal estimate as nominally significant.

We can determine if the causal estimates of the mQTL analyses seem to represent metabolism by ensuring that the causal estimate is positive, which would represent a chemical reaction between a substrate and a product in equilibrium conditions. We test for this “*le Chatelliers principle*” By taking the proportion of positive causal estimates compared to the total number of Bonferroni significant MR estimates? (**Fig. 4e-h**) (**Supplementary Text**).

#### Reference set of metabolite reactions

To benchmark the *cis* genetic methods used in this publication, we define a ground truth set of reactions. Real reactions are defined when the substrate ‘causes’ changes in the levels of the product. Unfortunately, it is difficult to define false causal links, as it is almost impossible to prove that two metabolites are not in a reaction together. On top of that, it could be that a metabolite is understudied and therefore a potential reaction is simply not known. Our approach defines negative metabolite combinations based on distance in the graph of “ground truth” reactions. If the minimum distance (counted as reactions) between metabolites is a certain number of steps or more, we consider the combination as a ground truth null reaction, as it is unlikely that any statistical method will have the power to pick up a signal after a certain number of reactions. We do not consider metabolite combinations a false causal link if there is no path possible between them in the extended graph for two reasons. First, we reduce the chance of an understudied metabolite being considered non-causal as they are present in the metabolite network that we test. Secondly, this approach ensures that the exposures studied are used both for true causal link and false causal links datasets, making the analysis less dependent on which associated regions are used, as they are the same or similar between all sets.

### Meta-analyzing multiple associated regions

When there are multiple associated gene regions available for a metabolite, it becomes possible to meta-analyze regions. We meta-analyze regions by taking the weighted mean 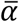 (and standard error 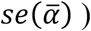of all associated regions together:

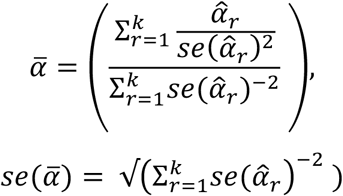

Where *k* are all the associated regions found in the initial clumping step, 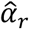 is the regional estimate with its associated standard error: 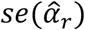.

### Analysis of complex traits

#### MR detection rates and false positive rates canonical causality

Next to metabolites, we turn to canonical causal relationships between complex traits. We perform regional estimates to understand the per region false positive rate and detection rate of MR methods (**Fig. 5a-b**). For each regional causal estimate we determine if it is nominally significant (0.05) and report the proportion of causal.

#### Heterogeneity of causal estimates

We meta-analyze the causal estimates of canonical causality in the same way as the metabolites (**Methods**). We estimate Cochran’s Q statistics across all these associated regions by taking the sum of the Z score deviation of a regional estimate with the meta-analyzed estimate for each trait pair (**Fig. 5c**).

#### Understanding the causal relationship between HDL-cholesterol and coronary artery disease

MR methods usually have trouble to correctly identify the non-causal relationship between HDL-C and CAD. This is due to the shared genetic regulation of HDL-C and LDL-C, where LDL-C is the causal biomarker. We use a heuristic to understand when MR methods stop identifying the causal relationship between HDL-C and CAD. We do this by iteratively selecting associated regions that are at least a specific number of base pairs away from other lipid regions and reporting their meta-analyzed P value (**Fig. 5d**).

### Analysis of gene expression and cell types

We used cell type definitions and their marker genes to identify if MR methods correctly identify the causal direction between cell types and their marker genes. Marker genes were taken from the Azimuth PBMC cell type reference (http://www.mhi-humangenetics.org/en/resources/) (*47*), which is typically used to identify cell types from single cell RNA sequencing experiments. We can identify marker genes for 10 out of 15 cell types. For each cell type composition / marker gene combination (41 in total), we perform bidirectional MR between cell types and their respective marker genes, where we assume that the marker genes are the cause of the cell type and not vice versa (**Data S14**) (**Data S16**) (**Fig. 5e**). We combine all cell type – marker gene causal relationships together and use P values to determine discriminative ability in terms of AUC (**Fig. 5f**).

## Supplementary Text

### The assumptions underlying MR and analogies to a randomized control trial (RCT)

Mendelian randomization (MR) is a statistical technique that can identify causal relationships from observational data. MR identifies a causal relationship in a process that can be related to a randomized control trial (RCT), with violations of the assumptions paralleling caveats in the RCT design (*69*). At conception, all humans randomly receive alleles from their parents. Some of these alleles have effects on human traits which allows us to group individuals into ‘genetic treatment’ groups based on the genetic variants they carry. MR identifies a causal relationship if the genetic treatment of the risk factor of interest is proportional to the genetics of the outcome trait under investigation. After all, if A is causal to B, all the genetics of trait A should also be visible in the genetics of trait B.

MR is valid under three main assumptions that can also be seen as incorrect applications of RCTs:

i. The relevance assumption states that the variants selected for the genetic treatment, also known as instrumental variables, should be relevant to the risk factor under investigation (**Fig. 1a**). In an RCT, this assumption would be violated when the treatment group receives a treatment that has no effect on the risk factor of interest. In MR, genetic variants are usually selected if strongly associated with the risk factor to avoid violating this assumption.
ii. The independence assumption states that the genetic variants under investigation should be independent from confounding with the outcome (**Fig. 1a**). In an RCT, this assumption would be violated when there is incorrect randomization. For instance, putting all the smokers in the control group will likely influence trial outcome. In MR this is difficult to test for, as unobserved population stratification can be a source of this violation (*70*). Recent evidence suggests that the independence assumption could be violated in the presence of population stratification, assortative mating and dynastic effects. This assumption is difficult to explore in the absence of family data.
iii. The exclusion restriction (also known as horizontal pleiotropy) states that the genetic variant should only affect the outcome through paths that are completely mediated by the risk factor (**Fig. 1a**). In an RCT, this assumption would be violated when the treatment contains some form of contamination that also affects the outcome. In this case it is impossible to discern if the treatment has an effect or if it is due to the contamination. In MR, it is difficult to test for the exclusion restriction as the genetic variants selected as instrumental variables that are used for the ‘genetic treatment’ can have unknown effects that are also causal to the outcome.

### Le Chateliers principle and MR on metabolism

*Le Chateliers* principle states that an increase in concentrations of a substrate also increases the concentrations of a product in a system that is in equilibrium. When applying Mendelian randomization (MR) on metabolites that are in a chemical reaction with one another, we expect that the causal estimate will be positive: an increase in the substrate (exposure in MR) will also increase the product (outcome in MR). One important caveat is that this positive effect is not what is initially expected when the genetic variants that are used for MR causally affect the enzyme that catalyzes a reaction. Interestingly, such a variant will have an effect on both the substrate concentrations and the product concentrations. If the genetic variant reduces the conversion efficiency of the enzyme, this will lead to an increase in the substrate concentrations and a decrease in the product concentrations as less substrate is converted. The MR effect should then seemingly be negative. Interestingly, this case is form of a violation of the exclusion restriction (no horizontal pleiotropy assumption) in an MR framework. The genetic variant that is used will have an effect on the outcome (the concentration of the product in a reaction) that is not directly mediated by the exposure (substrate). On top of this, this also violates Le Chateliers principle, as the equilibrium of the reaction is changed between carriers of the variant compared to non-carriers. So, even though a positive MR estimate is seemingly not expected when a genetic variant affects the enzyme catalyzing a reaction, due to a violation of the exclusion restriction as well as le Chatteliers principle not holding, the MR estimate should still remain positive in these cases.

## Derivation of the MR-link 2 likelihood function

Let traits *X* and *Y* represent two human traits, where *X* has a causal effect on *Y*, its size denoted by *α*. Moreover, let 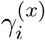 and 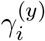 represent the multivariable effects of SNP *i* on *X* and *Y*. The vectorized version of these per SNP effects (representing the effect of all SNPs) are analogously denoted ***γ***(^*x*^), ***γ***(^*y*^), respectively. The underlying statistical model is as follows:

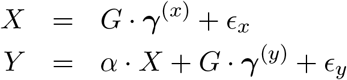

with *ϵ*_*x*_ and *ϵ*_*y*_ normally distributed errors. We also assume that the multivariate effect sizes come from a normal distribution, i.e. 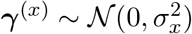 and 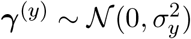. Subsequently, let 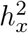 and 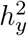 denote the direct local heritabilities of *X* and *Y*, thus 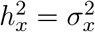 and 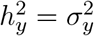.Let us assume that we have now data available for *X* in a sample of size *n*_*x*_ and for *Y* in a sample of *n*_*y*_. For simplicity we assume that both *X* and *Y* and the genotype data for each SNP have zero mean and unit variance (across the samples). This determines the error variances as follows: 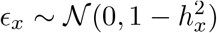 and 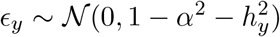. By multiplying both sides by *G*^*′*^ and divide the first equation by *n*_*x*_ and the second equation by *n*_*y*_ we have

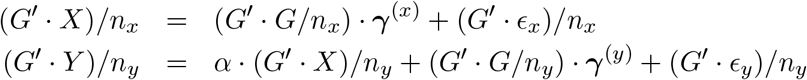

Let us assume that summary statistics (standardised marginal effect size estimates based on the previously stated sample sizes) are available for all *m* SNPs at a genomic region for trait *X* and *Y*, denoted by 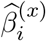 and 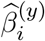 and the corresponding collection of these values in a vector form is 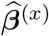 and 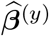. Furthermore, let *C* be the local *m × m* LD matrix. With these notations, we are in position to reformulate the equations as

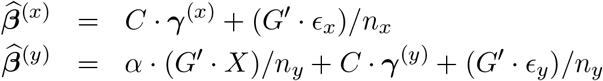

Substituting into *X* the first equation (although estimated in a difference sample (of size *n*_*y*_)) gives

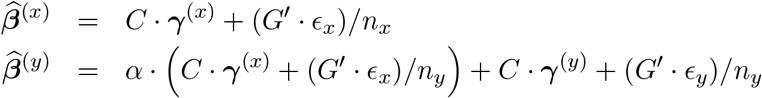

Denoting 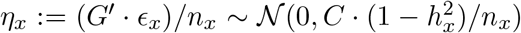 and 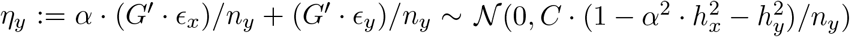,we have

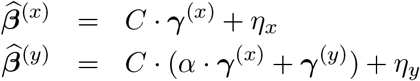

Assuming that the marginal effect estimates come from non-overlapping samples (i.e. their errors are uncorrelated), the likelihood function can be written

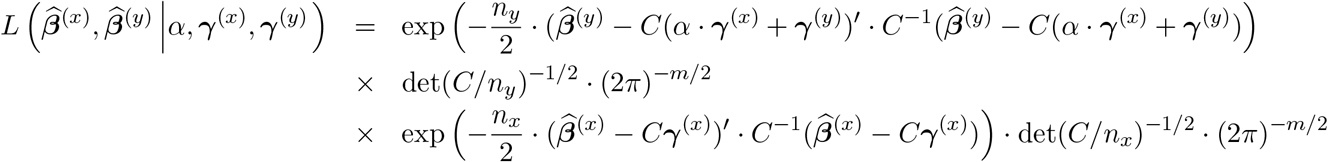

If we set the following priors for ***γ***(^*x*^), ***γ***(^*y*^), reflecting the InSIDE assumption (covariance being zero):

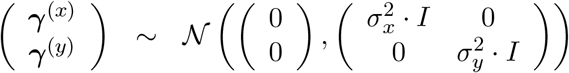

We can write

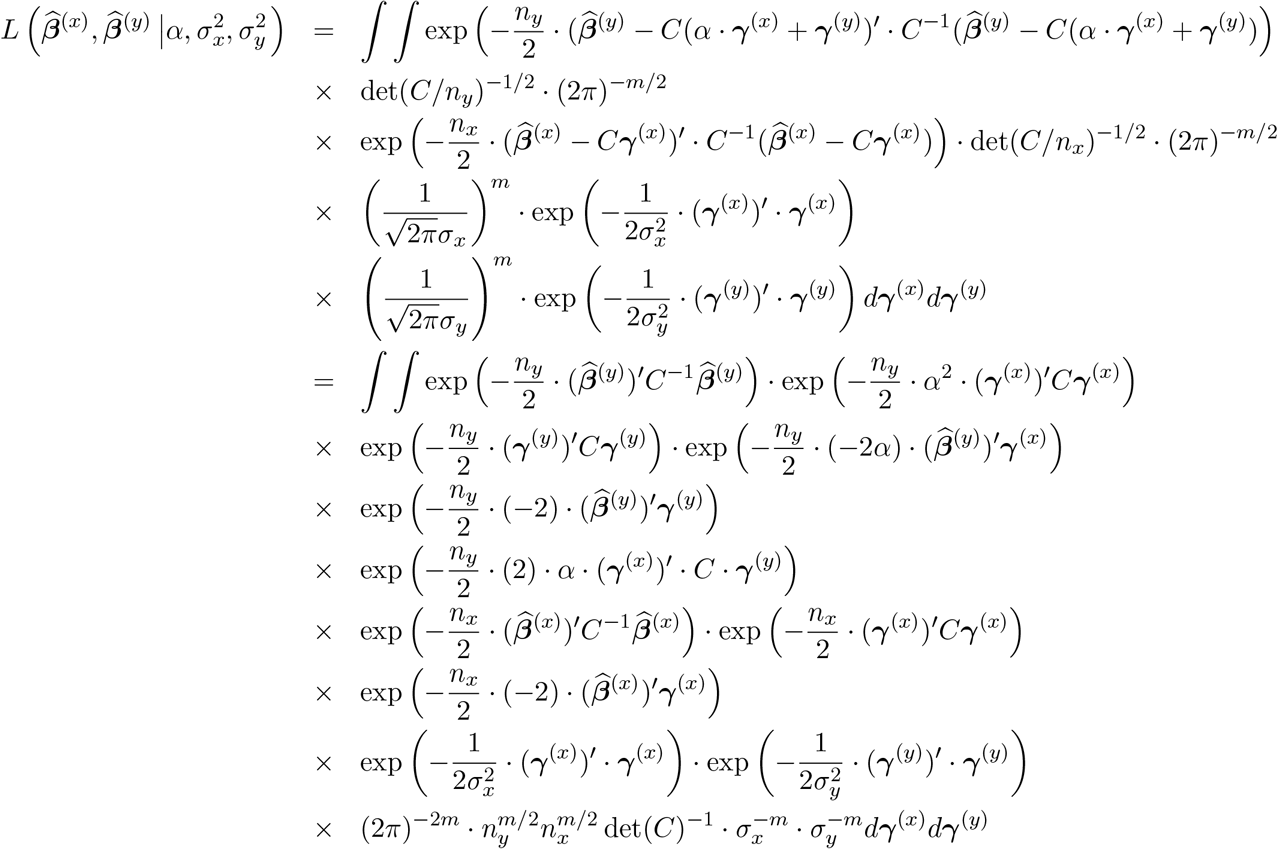

Next we complete the squares for ***γ***(^*x*^) and ***γ***(^*y*^)

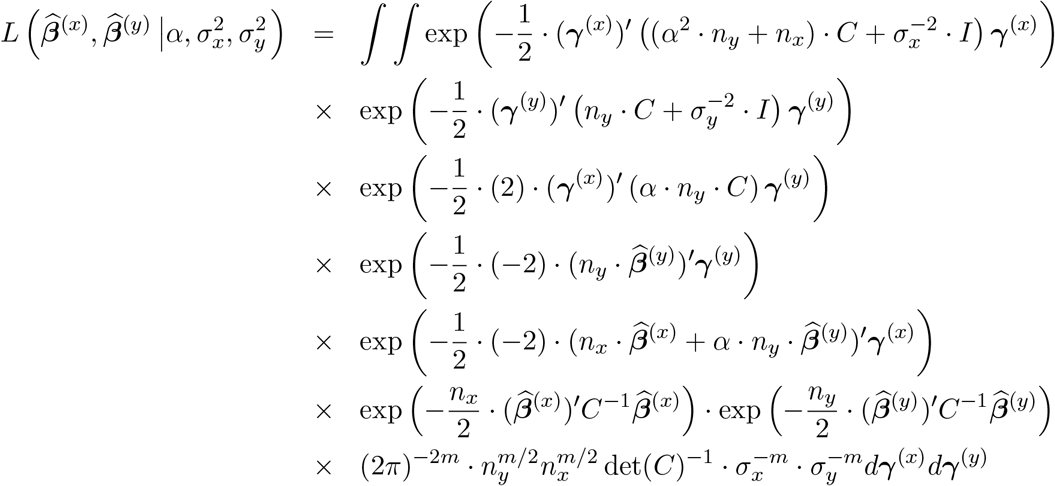

By introducing the following notations

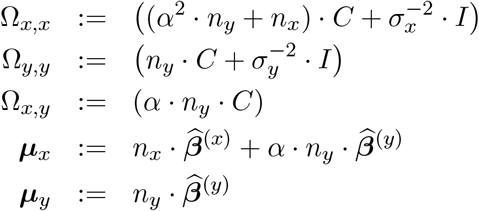

and using these notations further shorthand can be used

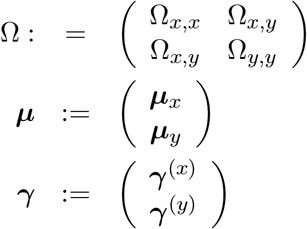

This allows us to turn the likelihood function into

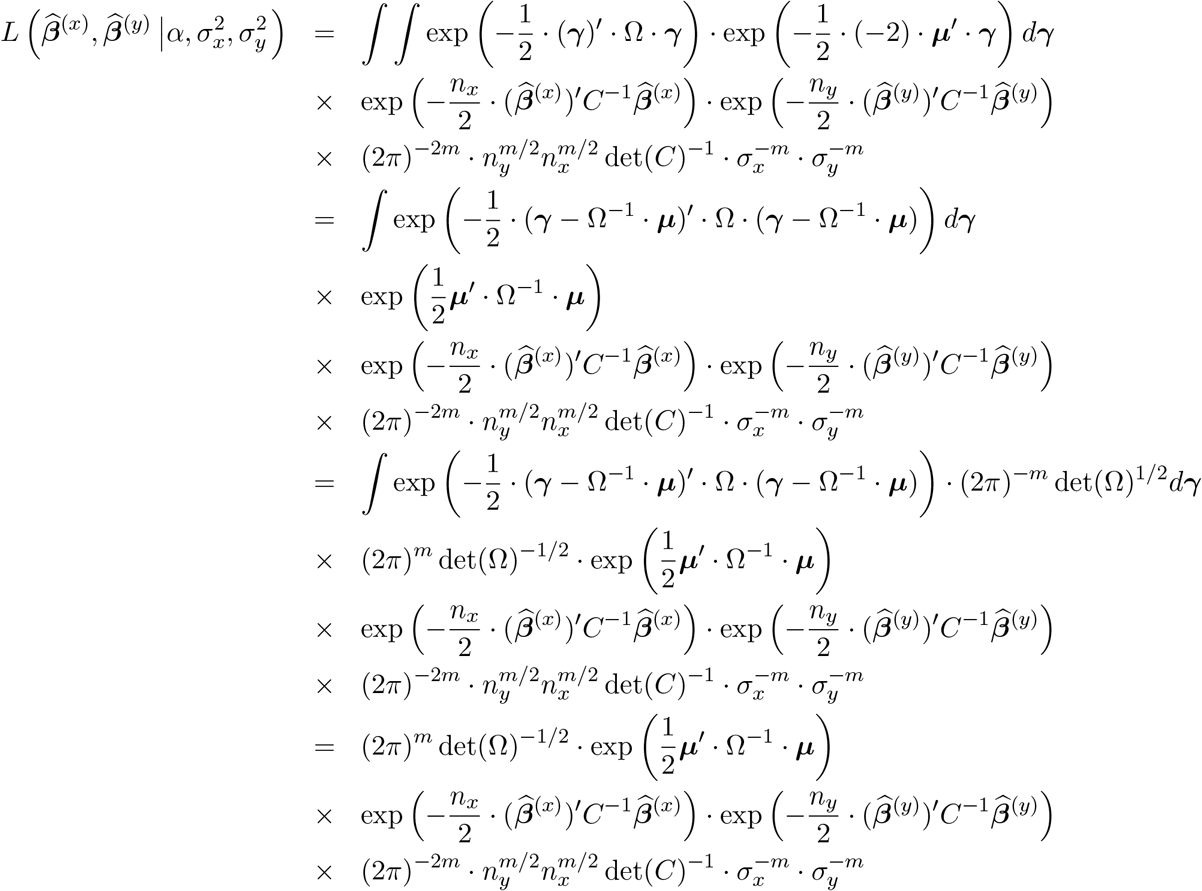

Let us replace *C* with its singular value decomposition *U ·* Λ *· U*^*′*^ with the *i*th diagonal element of Λ being *λ*_*i*_. In addition, let’s introduce the notation ***ν***_*x*_ := *U*^*′*^ *·* ***μ***_*x*_ and ***ν***_*y*_ := *U*^*′*^ *·* ***μ***_*y*_ and analogously, 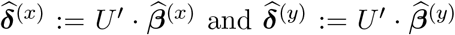. This allows the reformulation of Ω_*x*,*x*_, Ω_*y*,*y*_ and Ω_*x*,*y*_ as follows:

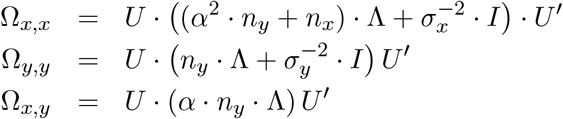

Using the formula for block matrix determinant, det(Ω) = det(Ω_*x*,*x*_ − Ω_*x*,*y*_Ω^−1^ Ω_*y*,*x*_) *·* det(Ω_*y*,*y*_), we have

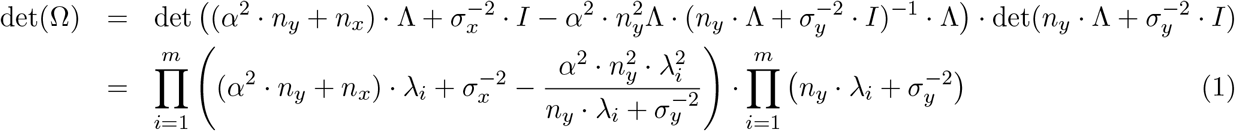

This time the formula of the block matrix inverse allows us the calculation of the block elements of the inverse of Ω:

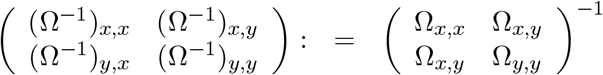

The first block can b e written as

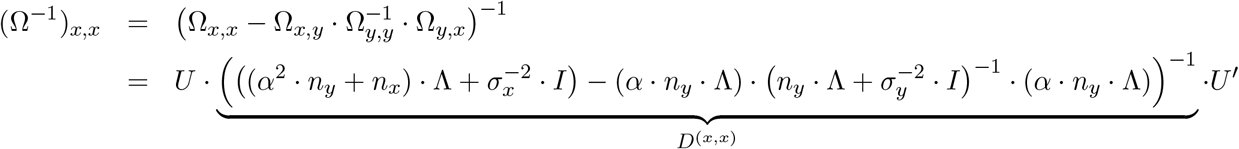

where *D*(^*x*,*x*^) is a diagonal matrix with elements

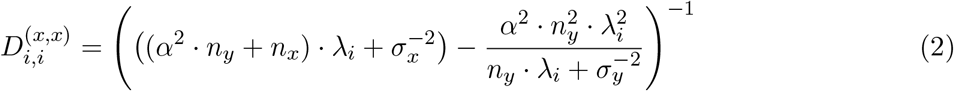

The second block can be written as

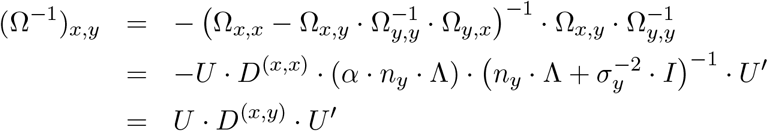

where *D*(^*x*,*y*^) is a diagonal matrix with elements

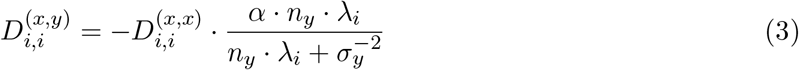

Finally, the last block is of the form

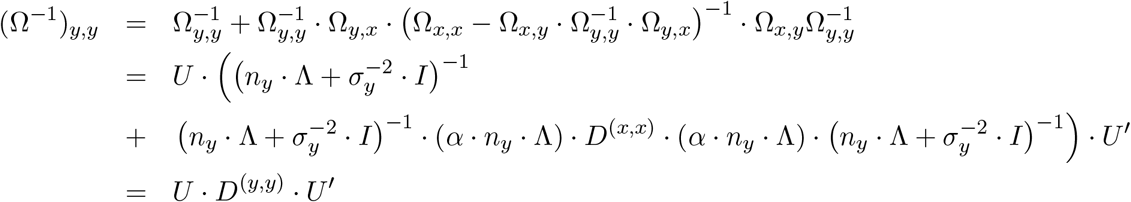

where *D*(^*y*,*y*^) is a diagonal matrix with elements

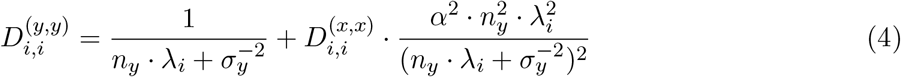

The bilinear product in the likelihood function can be written as

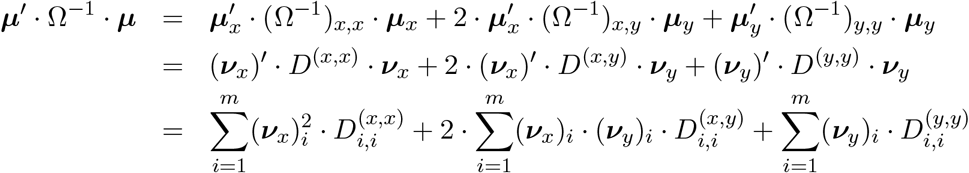

Analogously, we have

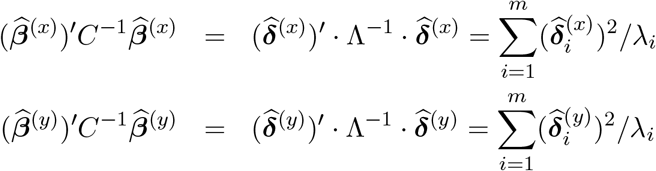

This puts us in position to simplify the log-likelihood function to

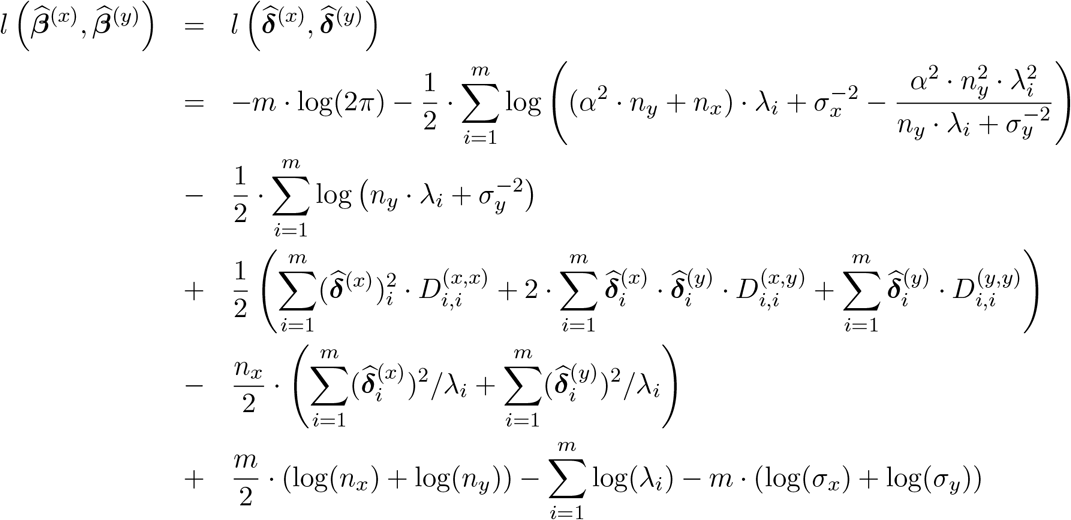

**eQTLGen Consortium – Author information**

**Toni Boltz**^**1**^, **Dorret Boomsma**^**2**^, **Andrew Brown**^**3**^, **Evans Cheruiyot**^**4**^, **Emma E. Davenport**^**5**^, **Theo Dupuis**^**3**^, **Tõnu Esko**^**6**^, **Aiman Farzeen**^**7**,**8**^, **Lude Franke**^**9**^, **Tim Frayling**^**10**^, **Greg Gibson**^**11**^, **Marleen van. Greevenbroek**^**12**^, **Michael Inouye**^**13**,**14**,**15**,**16**,**17**,**13**^, **Mika Kähönen**^**18**,**19**^, **Viktorija Kukushkina**^**6**^, **Sandra Lapinska**^**20**^, **Terho Lehtimäki**^**21**,**18**,**22**^, **Reedik Mägi**^**6**^, **Allan McRae**^**23**^, **Joyce van. Meurs**^**24**^, **Lili Milani**^**6**^, **Binisha Hamal Mishra**^**21**,**18**,**22**^, **Grant W. Montgomery**^**25**^, **Martina Müller-Nurasyid**^**26**^, **Sini Nagpal**^**11**^, **Matthias Nauck**^**27**,**28**^, **Roel Ophoff**^**20**,**1**,**29**^, **Bogdan Pasaniuc**^**20**,**30**,**1**,**31**^, **Dirk S. Paul**^**13**,**14**,**32**^, **Elodie Persyn**^**13**,**14**,**33**^, **Holger Prokisch**^**7**,**8**^, **Olli T. Raitakari**^**34**,**35**,**36**,**37**^, **Emma Raitoharju**^**38**,**39**^, **Jansen Rick**^**40**,**41**,**42**^, **Eline Slagboom**^**43**^, **Alexander Teumer**^**28**,**44**,**45**^, **Alex Tokolyi**^**5**^, **Jan Veldink**^**46**^, **Joost Verlouw**^**47**^, **Peter M. Visscher**^**48**^, **Uwe Völker**^**28**,**49**^, **Ur mo Võsa**^**6**^, **Robert Warmerdam**^**9**^, **Stefan Weiss**^**50**,**51**^, **Harm-Jan Westra**^**9**^, **Andrew Wood**^**10**^, **Manke Xie**^**11**,^

**Author list is ordered alphabetically**.

1. Department of Human Genetics, David Geffen School of Medicine, University of California Los Angeles, Los Angeles, CA, USA
2. Department of Biological Psychology, Vrije Universiteit Amsterdam, Amsterdam, The Netherlands and Amsterdam Public Health Research Institute, Amsterdam, The Netherlands.
3. Population Health and Genomics, Ninewells Hospital and Medical School, University of Dundee, Dundee, DD1 9SY, United Kingdom
4. School of Applied Systems Biology, La Trobe University, Bundoora, VIC, Australia.
5. Wellcome Sanger Institute, Wellcome Genome Campus, Hinxton, UK
6. Estonian Genome Centre, Institute of Genomics, University of Tartu, Tartu, Estonia
7. School of Medicine, Institute of Human Genetics, Klinikum rechts der Isar, Technical University of Munich, Munich, Germany
8. Institute of Neurogenomics, Computational Health Centre, Helmholtz Zentrum München, Neuherberg, Germany
9. Department of Genetics, University of Groningen, University Medical Centre Groningen, Groningen, The Netherlands
10. Genetics of Complex Traits, University of Exeter Medical School, University of Exeter, Exeter, UK
11. Center for Integrative Genomics, Georgia Institute of Technology, Atlanta, GA.
12. CARIM, Maastricht University
13. British Heart Foundation Cardiovascular Epidemiology Unit, Department of Public Health and Primary Care, University of Cambridge, Cambridge, UK
14. Victor Phillip Dahdaleh Heart and Lung Research Institute, University of Cambridge, Cambridge, UK
15. Cambridge Baker Systems Genomics Initiative, Department of Public Health and Primary Care, University of Cambridge, Cambridge, UK
16. Cambridge Baker Systems Genomics Initiative, Baker Heart and Diabetes Institute, Melbourne, VIC, Australia
17. Health Data Research UK Cambridge, Wellcome Genome Campus and University of Cambridge, Cambridge, UK
18. Finnish Cardiovascular Research Center Tampere, Faculty of Medicine and Health Technology, Tampere University, Tampere, Finland
19. Department of Clinical Physiology, Tampere University Hospital, Tampere Finland
20. Bioinformatics Interdepartmental Program, University of California Los Angeles, Los Angeles, CA, USA
21. Department of Clinical Chemistry, Faculty of Medicine and Health Technology, Tampere University, Tampere, Finland
22. Department of Clinical Chemistry, Fimlab Laboratories, Tampere, Finland
23. Institute for Molecular Bioscience, The University of Queensland, Brisbane, Australia
24. Department of Internal Medicine, Erasmus MC University Medical Center Rotterdam, Rotterdam, the Netherlands
25. Institute for Molecular Bioscience, The University of Queensland, Carmody Rd, Brisbane, QLD, 4067, Australia
26. Institute of Genetic Epidemiology, Helmholtz Zentrum München - German Research Center for Environmental Health, Neuherberg, Germany
27. Institute of Clinical Chemistry and Laboratory Medicine, University Medicine Greifswald, Greifswald, Germany
28. DZHK (German Centre for Cardiovascular Research), Partner Site Greifswald, University Medicine, Greifswald, Germany
29. Center for Neurobehavioral Genetics, Semel Institute for Neuroscience and Human Behavior, David Geffen School of Medicine, University of California Los Angeles, Los Angeles, USA
30. Department of Computational Medicine, David Geffen School of Medicine, University of California Los Angeles, Los Angeles, CA, USA
31. Department of Pathology and Laboratory Medicine, David Geffen School of Medicine, University of California Los Angeles, Los Angeles, CA, USA
32. Centre for Genomics Research, Discovery Sciences, BioPharmaceuticals R&D, AstraZeneca, Cambridge, UK
33. Cambridge Baker Systems Genomics Initiative, Department of Public Health and Primary Care, University of Cambridge, UK
34. Research centre of Applied and Preventive Cardiovascular Medicine, University of Turku, Turku, Finland
35. Department of Clinical Physiology and Nuclear Medicine, Turku University Hospital, Turku, Finland
36. Centre for Population Health Research, University of Turku and Turku University Hospital, Turku, Finland
37. InFLAMES Research Flagship, University of Turku, Turku, Finland
38. Molecular Epidemiology, Faculty of Medicine and Health Technology, Tampere University, Tampere, Finland
39. Tampere University Hospital, Tampere, Finland
40. Amsterdam UMC location Vrije Universiteit Amsterdam, Department of Psychiatry, Amsterdam, the Netherlands Amsterdam, the Netherlands.
41. Amsterdam Public Health, Mental Health Program, Amsterdam, the Netherlands
42. Amsterdam Neuroscience, Mood, Anxiety, Psychosis, Sleep & Stress Program, Amsterdam, the Netherlands.
43. Section of Molecular Epidemiology, Department of Biomedical Data Sciences, Leiden University Medical Center, PO Box 9600, Post-zone S-05-P, 2300 RC, Leiden, The Netherlands.
44. Institute for Community Medicine, University Medicine Greifswald, Greifswald, Germany
45. Department of Psychiatry and Psychotherapy, University Medicine Greifswald, Greifswald, Germany
46. Department of Neurology, Brain Center Rudolf Magnus, University Medical Center Utrecht, Utrecht, The Netherlands
47. Department of Internal Medicine, Erasmus University Medical Center, Rotterdam, The Netherlands
48. Institute for Molecular Bioscience, The University of Queensland, Brisbane, Queensland, Australia
49. Interfaculty Institute of Genetics and Functional Genomics, Department Functional Genomics, University Medicine Greifswald, Greifswald, Germany
50. Interfaculty Institute for Genetics and Functional Genomics, University Medicine Greifswald and University of Greifswald, Greifswald, Germany
51. German Center for Cardiovascular Research, Greifswald, Germany

### eQTLGen cohort information

The details for the majority of eQTLGen cohorts used in the preliminary meta-analysis freeze are detailed in the Võsa & Claringbould et al., 2021, and corresponding original publications. Below we provide the details of additional INTERVAL cohort which was not part of Võsa & Claringbould et al., 2021. (*21*)

The eQTLGen phase II research activities involving Estonian Biobank participant data (two EstBB cohorts) have been carried out under the ethical approval nr. 1.1-12/655 and its extension 1.1-12/490 by the Estonian Committee on Bioethics and Human Research (Estonian Ministry of Social Affairs), using data according to release application number S54 from the Estonian Biobank.

## INTERVAL

The INTERVAL study is a prospective cohort study of approximately 50,000 participants nested within a randomized trial of varying blood donation intervals (*71, 72*). Between 2012 and 2014, blood donors aged 18 years and older were recruited at 25 centers of England’s National Health Service Blood and Transplant (NHSBT). Participants were generally in good health as blood donation criteria exclude individuals with a history of major diseases (e.g. myocardial infarction, stroke, cancer, HIV, and hepatitis B or C) and who have had a recent illness or infection.

Participants completed an online questionnaire comprising questions on demographic characteristics (e.g. age, sex, ethnicity), lifestyle (e.g. alcohol and tobacco consumption), self-reported height and weight, diet and use of medications. All participants gave informed consent before joining the study and the National Research Ethics Service approved this study (11/EE/0538).

### Blood collection

Blood samples were collected from all INTERVAL participants at baseline and also from ∼60% of participants approximately 24 months after baseline. For a subset of ∼5,000 participants at the 24-month time point, an aliquot of 3 ml of whole blood was collected in Tempus Blood RNA Tubes (ThermoFisher Scientific), following the manufacturer’s instructions, and then transferred at ambient temperature to the UK Biocentre (Stockport, UK). Samples were stored at -80°C until use.

### RNA extraction

RNA extraction was performed by QIAGEN Genomic Services using QIAGEN’s proprietary silica technology. The quality control of the extracted RNA was performed by spectrophotometric measurement on an Infinite 200 Microplate Reader (Tecan). RNA Integrity Number (RIN) values were determined using a TapeStation 4200 system (Agilent), following the manufacturer’s protocol. Samples with a concentration <20 ng/μl and a RIN value <4 were excluded from further analyses.

### Automated RNA-seq library preparation

Samples were quantified with a QuantiFluor RNA System (Promega) using a Mosquito LV liquid handling platform (SPT Labtech), Bravo automation system (Agilent) and FLUOstar Omega plate reader (BMG Labtech), and then cherry-picked to 200 ng in 50 μl (= 4 ng/μl) using a liquid handling platform (Tecan Freedom EVO). Next, mRNA was isolated using a NEBNext Poly(A) mRNA Magnetic Isolation Module (NEB) and then re-suspended in nuclease-free water. Globin depletion was performed using a KAPA RiboErase Globin Kit (Roche). RNA library preparation was done using a NEBNext Ultra II DNA Library Prep Kit for Illumina (NEB) on a Bravo NGS workstation automation system (Agilent). PCR was performed using a KapaHiFi HotStart ReadyMix (Roche) and unique dual-indexed tag barcodes on a Bravo NGS workstation automation system (Agilent). We applied the following PCR programme: 45 sec at 98°C, 14 cycles of 15 sec at 98°C, 30 sec at 65°C and 30 sec at 72°C, followed by 60 sec at 72°C. Using a Zephyr liquid handling platform (PerkinElmer), PCR products were purified using AMPure XP SPRI beads (Agencourt) at a 0.8:1 bead:sample ratio and then eluted in 20 μl of Elution Buffer (QIAGEN). RNA-seq libraries were quantified with an AccuClear Ultra High Sensitivity dsDNA Quantitation Kit (Biotium) using a Mosquito LV liquid handling platform (SPT Labtech), Bravo automation system (Agilent) and FLUOstar Omega plate reader (BMG Labtech). Then, libraries were pooled up to 95-plex in equimolar amounts on a Biomek NX-8 liquid handling platform (Beckman Coulter), quantified using a High Sensitivity DNA Kit on a 2100 Bioanalyzer (Agilent), and then normalized to 2.8 nM prior to sequencing.

### RNA sequencing and data pre-processing

Samples were sequenced using 75 bp paired-end sequencing reads (reverse stranded) on a NovaSeq 6000 system (S4 flow cell, Xp workflow; Illumina). The sequencing data were de-plexed into separate CRAM files for each library in a lane. Adapters that had been hard-clipped prior to alignment were reinserted as soft-clipped post alignment, and duplicated fragments were marked in the CRAM files. The data pre-processing, including sequence QC, and STAR and alignments was performed with the Nextflow pipeline publicly available at https://github.com/wtsi-hgi/nextflow-pipelines/blob/rna_seq_interval_5591/pipelines/rna_seq.nf, including the specific aligner parameters. We assessed the sequence data quality using FastQC v0.11.8. Samples mismatched between RNA-seq and genotyping data within the cohort were identified using QTLtools MBV v1.2 (*73*). Reads were aligned to the GRCh38 human reference genome (Ensembl GTF annotation v99) using STAR v2.7.3a (*74*). The STAR index was built against GRCh38 Ensembl GTF v99 using the option -sjdbOverhang 75. STAR was run in a two-pass setup with standard ENCODE options to increase mapping accuracy: (i) a first alignment step of all samples was used to discover novel splice junctions; (ii) splice junctions of all samples from the first step were collected and merged into a single list; (iii) a second step realigned all samples using the merged splice junctions list as input. We used featureCounts v2.0.0 (*75*) to obtain a count matrix.

### Gene expression quantification

The raw gene-level count data contained N=60,676 genes across N=4,778 individuals with 2.03–95.55 million uniquely mapped reads (median: ∼24 million). Sequencing was performed across 15 batches.

### Quality control of gene expression data

We filtered samples of poor quality by removing samples with a read depth below 10 million uniquely mapped reads. A relatedness matrix was obtained using the PLINK v1.9 (*54*) -make-rel ‘square’ command on pruned genotype data, and a cut-off threshold of 0.1 was used to define related individuals. For each pair of related individuals, one individual was arbitrarily removed. After filtering, the gene expression dataset included 4,731 individuals. We retained 60,580 genes located on autosomal and sex chromosomes. Then, the raw expression matrix was automatically processed with eQTLGen pipelines.

### Genotyping data

In brief, DNA extracted from buffy coat samples collected from INTERVAL participants at the study baseline was used to assay approximately 830,000 variants on the Affymetrix Axiom UK Biobank genotyping array (*76*). Genotyping and sample QC were performed as previously described (*76*). Next, genotype data was automatically quality-controlled, pre-phased and imputed by eQTLGen pipelines.

## Data availability

The INTERVAL study data used in this paper are available to bona fide researchers from. The data access policy for the data is available at http://www.donorhealth-btru.nihr.ac.uk/project/bioresource. The generated RNA-sequencing data have been deposited at the European Genome-phenome Archive (EGA) under the accession number EGAD00001008015.

## Acknowledgements

Participants in the INTERVAL randomized controlled trial were recruited with the active collaboration of NHS Blood and Transplant England (https://www.nhsbt.nhs.uk/), which has supported field work and other elements of the trial. DNA extraction and genotyping were co-funded by the National Institute for Health and Care Research (NIHR), the NIHR BioResource (https://bioresource.nihr.ac.uk/) and the NIHR Cambridge Biomedical Research Centre (BRC-1215-20014) [*]. RNA-seq was funded as part of an alliance between the University of Cambridge and the AstraZeneca Centre for Genomics Research, and by the NIHR Cambridge Biomedical Research Centre (BRC-1215-20014) [*]. The academic coordinating center for INTERVAL was supported by core funding from the NIHR Blood and Transplant Research Unit (BTRU) in Donor Health and Genomics (NIHR BTRU-2014-10024); NIHR BTRU in Donor Health and Behaviour (NIHR203337); UK Medical Research Council (MR/L003120/1); British Heart Foundation (SP/09/002; RG/13/13/30194; RG/18/13/33946); and NIHR Cambridge BRC (BRC-1215-20014; NIHR203312) [*]. A complete list of the investigators and contributors to the INTERVAL trial is provided in Di Angelantonio *et al*. (*71*) The academic coordinating center would like to thank blood donor center staff and blood donors for participating in the INTERVAL trial. This work was supported by Health Data Research UK, which is funded by the UK Medical Research Council, Engineering and Physical Sciences Research Council, Economic and Social Research Council, Department of Health and Social Care (England), Chief Scientist Office of the Scottish Government Health and Social Care Directorates, Health and Social Care Research and Development Division (Welsh Government), Public Health Agency (Northern Ireland), British Heart Foundation and Wellcome. *The views expressed are those of the authors and not necessarily those of the NIHR or the Department of Health and Social Care. The Wellcome Sanger Institute is supported by core funding from the Wellcome Trust (206194 and 220540/Z/20/A). We thank the Wellcome Sanger Institute’s Scientific Operations team for their contribution to sequencing data generation. For the purpose of Open Access, the authors have applied a CC BY public copyright licence to any Author Accepted Manuscript version arising from this submission. This work was supported by the Cambridge Service for Data Driven Discovery (CSD3) operated by the University of Cambridge Research Computing Service (https://www.csd3.cam.ac.uk/), provided by Dell EMC and Intel using Tier-2 funding from the Engineering and Physical Sciences Research Council (capital grant EP/P020259/1), and DiRAC funding from the Science and Technology Facilities Council (https://dirac.ac.uk/).

## Funding information

E.P. was funded by the EU/EFPIA Innovative Medicines Initiative Joint Undertaking BigData@Heart grant 116074 and is funded by the NIHR BTRU in Donor Health and Behaviour (NIHR203337) [*]. A.T. is supported by the Wellcome Trust (PhD studentship 222548/Z/21/Z). M.I. is supported by the Munz Chair of Cardiovascular Prediction and Prevention and the NIHR Cambridge Biomedical Research Centre (BRC-1215-20014; NIHR203312) [*]. M.I. is also supported by the UK Economic and Social Research Council (ES/T013192/1). M.I. is a trustee of the Public Health Genomics (PHG) Foundation, a member of the Scientific Advisory Board of Open Targets, and has a research collaboration with AstraZeneca that is unrelated to this study. D.S.P. is an employee and stockholder of AstraZeneca.

**Data S1**. Simulation results of MR-link-2. Detection rates for the two parameters MR-link-2 tests for (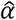 and 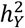 and the number of successful simulation runs. Columns are as follow: “simulated-alpha”: The *α* parameter simulated. “simulated-h^2_X”: The simulated exposure heritability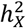. “simulated-h^2_Y”: The simulated outcome heritability 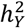 that would violate the exclusion restriction. “simulated-n_ref”: number of simulated individuals in the reference panel. When the number is 0 this means that the reference panel is simulated with full precision. “simulated-m_causal”: The number of causal SNPs simulated for the exposure *X* and the outcome *Y*. “simulated-min(r_causal)”: The minimum LD between causal SNPs. “simulated-max(r_causal)”: The maximum LD between causal SNPs. “number_of_mr-link2_estimates”: The number of successful estimates for MR-link-2. “MR-link_2_alpha_detection_rate”: The detection rate for the causal estimate of MR-link-2 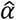 at P < 0.05. “MR-link_2_h^2_Y_detection_rate”: The detection rate for MR-link-2 for the pleiotropy parameter 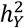.

**Data S2**. AUC comparisons in simulations of all the MR and coloc methods tested in this study. The columns are as follows: “simulated-alpha”: The *α* parameter simulated. “simulated-h^2_X”: The simulated exposure heritability 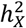. “simulated-h^2_Y”: The simulated outcome heritability 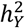 that would violate the exclusion restriction. “simulated-n_ref”: number of simulated individuals in the reference panel. When the number is 0 this means that the reference panel is simulated with full precision. “simulated-m_causal”: The number of causal SNPs simulated for the exposure *X* and the outcome *Y*. “simulated-min(r_causal)”: The minimum LD between causal SNPs. “simulated-max(r_causal)”: The maximum LD between causal SNPs. “method”: The method that makes the estimates. “number_of_null_estimates”: The number of estimates when not simulating a causal effect. “number_of_non_null_estimates”: The number of estimates when simulating a causal effect. “auc of the method”: The area under the receiver operator characteristic curve for the methods estimates.

**Data S3**. Regression of parameters of the simulation on the area under the receiver operator characteristic curve (AUC) of each method in this study. The columns are as follows: “explanatory”: the explanatory variable, the parameter of the simulation, except when the variable is ‘const’, then it is the intercept, “Coef.”: The coefficient of regression, “Std.Err.”: The standard error of the coefficient of regression, “t”: T statistic, “P>|t|”: p calue of the T statistic, “[0.025” and “0.975]” upper and lower confidence intervals of the coefficient, “method”: The AUC of the method as the explained variable

**Data S4**. Metabolites that are harmonized in this study, their identifiers in different databases, their metabolite quantitative trait locus (mQTL) study accession and indications where the metabolite has been used. The columns describe the harmonized name “harmonized_name”, Identification in different pathway databases: (“hmdb_id”, “inchikey” and “kegg_id”), the mQTL study accession “accession” and “study” and finally if and how the metabolite is used in analysis “used_as_outcome_in_metabolite_networks” indicates if the metabolite is found in the pathway databases and has a direct reaction with another metabolite. “used_as_exposure_in_metabolite_networks” indicates if the metabolite has an associated region from which it is possible to perform MR and finally “used_in_self_comparisons” is used to define if the metabolite is used in self-comparisons where an estimate is made based on the same metabolite measured in another mQTL study.

**Data S5**. Regional metabolite self-comparisons. Here we report the causal estimates (“alpha_estimates”, “se(alpha)”, “P(alpha)”) of different MR methods “method” across comparisons of the same metabolites (‘metabolite_name’, “hmdb_id”) measured in different studies (“exposure accession”, “outcome_accession”) across associated regions for the exposure (“region”). The column “included_in_bias_analysis” describes if the estimate is included in the bias analysis, as the P value is nominally significant (P < 0.05).

**Data S6**. Regional metabolite to metabolite estimates. Here we report the causal estimates (“alpha_estimates”, “se(alpha)”, “P(alpha)”) of different MR methods “method” across comparisons of different metabolites for an exposure (“exposure_name”, “exposure_hmdb_id”, “exposure accession”) and an outcome (“outcome_name”, “outcome_hmdb_id”, “outcome_accession”) for each associated exposure region “associated region”. The number of clumped variants in each region is denoted by “ivs in region”.

**Data S7**. Area under the receiver operator characteristic curves (AUCs) for each method’s (“method”) regional estimates evaluated independently. This is done for each pathway reference (“pwy”) and minimum distance the metabolites are away from each other (“distance”) in the extended graph. The AUC metric is performed on the respective number of “positives” and “negatives”.

**Data S8**. “precision” and “recall” for all the methods in both assessment of per locus independently and weighed together (“type”) across different pathway references “pwy”. These values are estimated for each individual method “method”.

**Data S9**. Area under the receiver operator characteristic curves (AUCs) for each method (“method”) when regional estimates are weighted together. This is done for each pathway reference (“pwy”) and minimum distance the metabolites are away from each other (“distance”). The AUC metric is performed on a number of “positives” and “negatives”.

**Data S10**. Metabolite to metabolite estimates, all regions of an exposure outcome combination weighted together. Here we report the causal estimates (“alpha_estimates”, “se(alpha)”, “P(alpha)”) of different MR methods “method” across comparisons of different metabolites for an exposure (“exposure_name”, “exposure_hmdb_id”, “exposure accession”) and an outcome (“outcome_name”, “outcome_hmdb_id”, “outcome_accession”). The number of regions weighted together is represented by “# Meta-analyzed regions”.

**Data S11**. The causal estimates all complex-complex trait combinations. Here we report the causal estimates (“Weighted alpha estimates”, “se(alpha)”, “P(alpha)”) of different MR methods “Method” across comparisons of different metabolites for an exposure (“exposure_name”,) and an outcome (“outcome_name”). The number meta-analyzed associated regions is denoted by ‘# of Meta-analyzed regions’.

**Data S12**. Detection rates at P < 0.05 per associated region (“Ratio of regions P < 0.05’) of a complex trait combination (“Exposure trait”, “Outcome trait”) that is considered causal or not causal (“Considered causal”), for each MR method tested “method”.

**Data S13**. Inverse variance weighted (across associated regions) analysis of MR estimates for each MR method tested in this study (“Method”) of HDL cholesterol on coronary artery disease, subsampling for associated regions that are at least a certain base-pair distance (“distance (bp)”) away from any region associated to LDL cholesterol, total cholesterol and non-HDL cholesterol associated regions. The number of regions that are retained are referenced by “n_regions_included”. Causal estimates are represented by “Weighted alpha estimate”, their P value (“P alpha”) and the meta-analyzed weights.

**Data S14**. Inverse variance weighted (across associated regions) analysis of cell types on gene expression and vice versa. Here we report the causal estimates (“Weighted alpha estimates”, “se(alpha)”, “P(alpha)”) of different MR methods “Method” across comparisons of different metabolites for an exposure (“exposure_name”,) and an outcome (“outcome_name”). The numbers of in regions analyzed is denoted by “# of Meta-analyzed regions”. We also report the Cochran’s Q heterogeneity statistic (“Cochrans Q statistic of meta-analysis”) for each combination and if the causal combination is considered causal. We consider the comparison causal if cell type composition differences influence their markergene expression.

**Data S15**. The summary statistics of all the complex traits that are used in this study, including the trait of interest “Complex trait”, the origin of the summary statistics file “Publication”, the maximum sample size for the trait and the doi identifier of the publication.

**Data S16**. Cell type (“Celltype”) and marker gene (“markergene”) reference adapted from the Azimuth reference. These cell type and markergene combinations are considered the true causal links that are used in the cell type analysis.

**Fig. S1.**
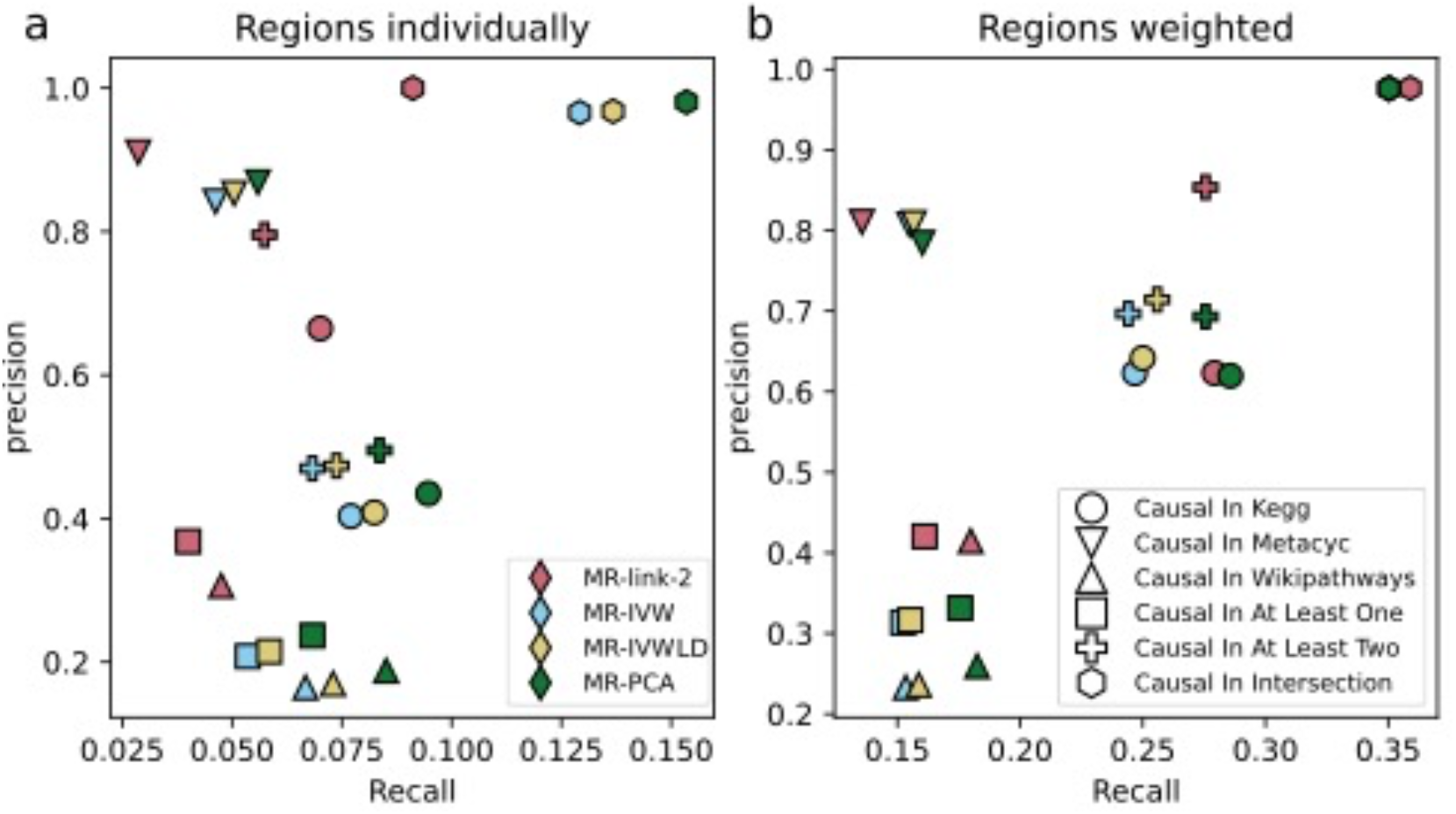
Precision and recall for the *cis* MR methods in this study. Each point is colored based on the respective method and the shape of each point represents the ground truth the method is tested on. (**a**) The precision and recall at Bonferroni significance (*P* < 2.3 ⋅ 10^−7^) when considering each region individually. (**b**) The precision and recall at Bonferroni significance (P < 1.0 ⋅ 10^−6^) when considering the inverse variance weighted estimate of each region together.

**Fig. S2.**
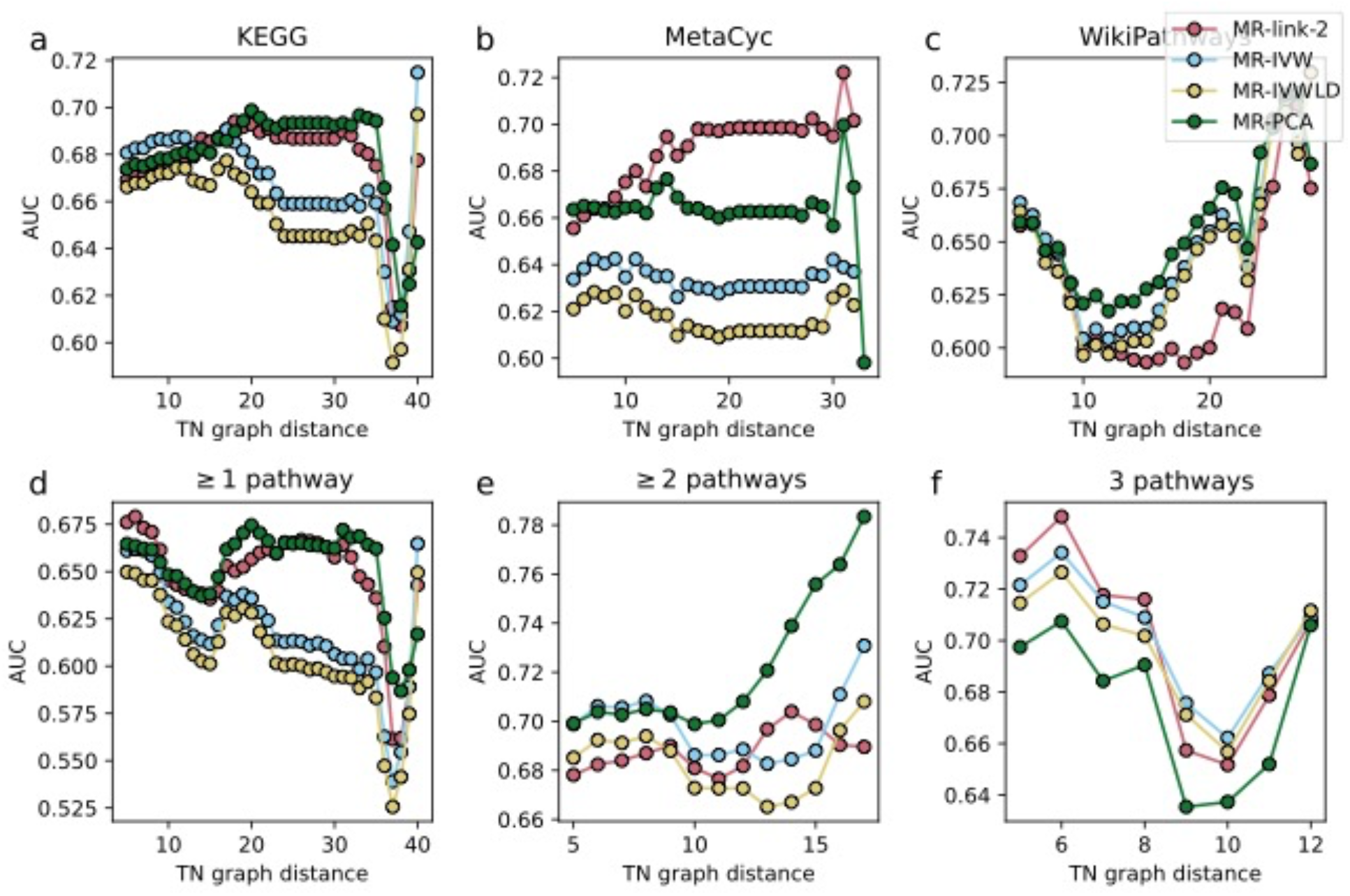
The area under the receiver operator characteristic curve (AUC) of the *cis* MR methods in this study when meta-analyzed together, benchmarked against different databases (**a**-**c**) and database combinations (**d**-**f**). Only showing comparisons when there are more than 10 negatives per positive definition (**Data S9**). (**a**) True causal links and false causal links from the KEGG pathway, (**b**) true causal links and false causal links from the MetaCyc pathway, (**c**) true causal links and false causal links from the WikiPathways pathway, (**d**) true causal links and false causal links that are present in any pathway definition, (**e**) true causal links and false causal links in at least two pathway definitions, (**f**) true causal links and false causal links that are shared in all pathways.

**Fig. S3.**
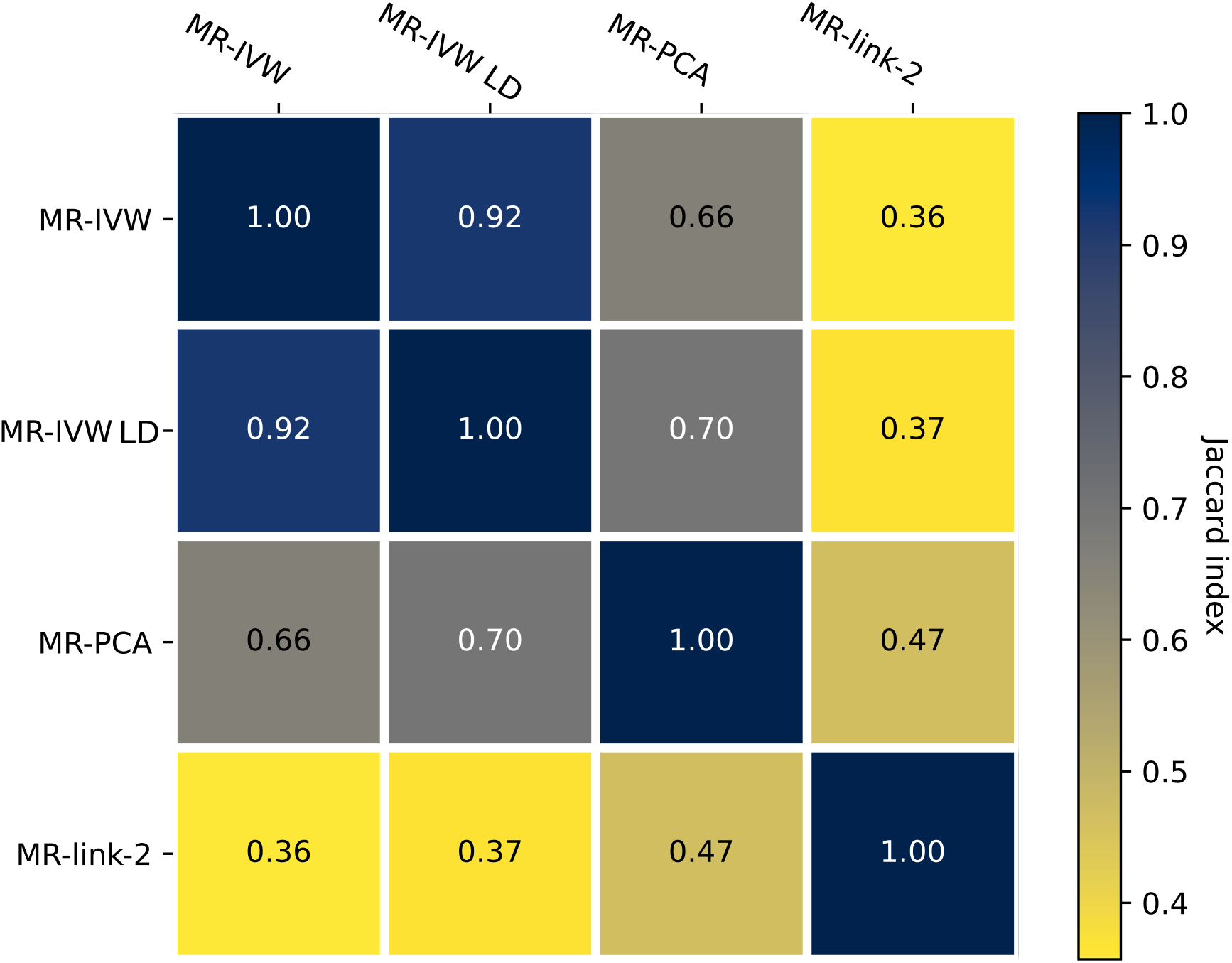
The Jaccard matrix of the (weighted across regions) Bonferroni significant (P < 9. 9 ⋅ 10^−6^) MR methods compared together. The Jaccard index is defined as sharing of the intersection of the set of causal relationships that two methods find divided by the union of the set of causal relationships.

**Fig. S4.**
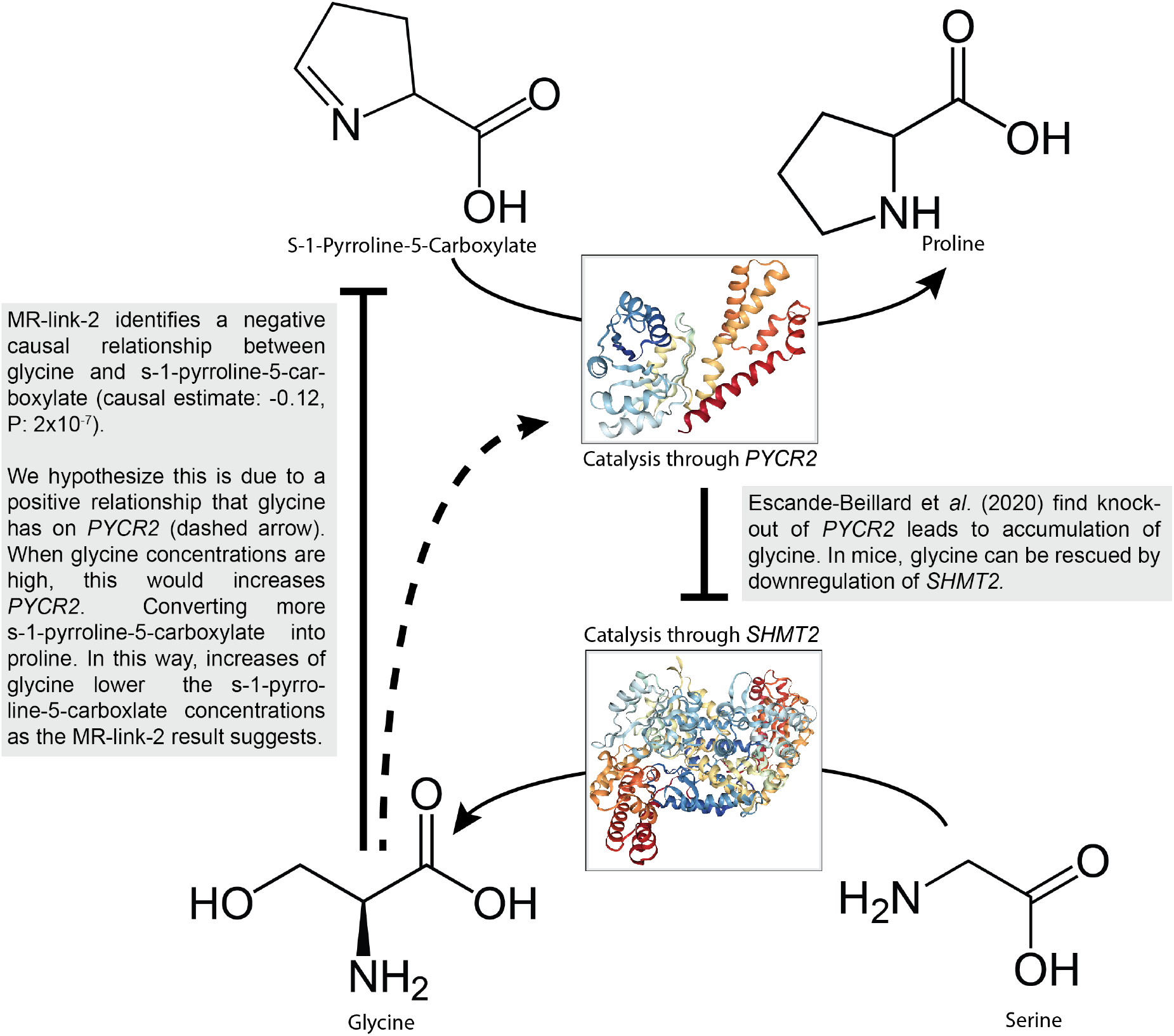
A visual overview of the causal relationship between glycine and s-1-pyrroline-5-carboxylate that MR-link-2 identifies. Here, the reaction catalyzed by the pyrroline-5-carboxylate reductase 2 (*PYCR2*) enzyme (image from PDB entry: 6LHM) converts s-1-pyrroline-5-carboxylate into proline. *PYCR2* inhibits the activity of the serine hydroxy-methyltransferase 2 (*SHMT2*) *enzyme* (image from PDB entry: 6QVL) that catalyzes the reaction between serine and glycine. This inhibiting reaction has been shown by Escande-Beillard et *al*. **(*40*)**. MR-link2 identifies a causal relationship between glycine and s-1-pyrroline-5-carboxylate 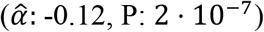 that we hypothesize is the result of a positive causal relationship between glycine and *PYCR2 (dashed line*)

## Notes

### Competing Interest Statement

The main authors of this study do not declare a competing interest.
The authors of the eQTLGen consortium declare the following competing interests:
B.M.P. serves on the Steering Committee for the Yale Open Data Access Project funded by Johnson & Johnson. This activity is unrelated to this work.
M.I. is a trustee of the Public Health Genomics (PHG) Foundation, a member of the Scientific Advisory Board of Open Targets, and has a research collaboration with AstraZeneca that is unrelated to this study.
D.S.P. is an employee and stockholder of AstraZeneca.
The other authors of the eQTLGen consortium do not declare competing interests.

### Funding Statement

This study was funded by the Swiss National Science Foundation (310030_189147, Zoltan Kutalik) and the Department of Computational Biology of the University of Lausanne (Zoltan Kutalik)

### Author Declarations

All data used in this study is publicly available except for newly generated data from the 2 cohorts in the eQTLgen consortium. The details for the previously available eQTLGen cohorts used in the preliminary meta-analysis freeze are detailed in the Vosa & Claringbould et al., 2021 publication, and corresponding original publications. Below we provide the details of additional INTERVAL cohort which was not part of Vosa & Claringbould et al., 2021. The eQTLGen phase II research activities involving Estonian Biobank participant data (two EstBB cohorts) have been carried out under the ethical approval nr. 1.1-12/655 and its extension 1.1-12/490 by the Estonian Committee on Bioethics and Human Research (Estonian Ministry of Social Affairs), using data according to release application number S54 from the Estonian Biobank. The INTERVAL study is a prospective cohort study of approximately 50,000 participants nested within a randomized trial of varying blood donation intervals. All participants gave informed consent before joining the study and the United Kingdom National Research Ethics Service approved this study (11/EE/0538).

